# A proteome-based classification of pediatric adrenocortical tumors links functional tumor states to clinical outcome and therapeutic vulnerabilities

**DOI:** 10.64898/2026.03.18.26348723

**Authors:** Rainer Claus, Andreas Metousis, Victoria E. Fincke, Marina Kunstreich, Stefan A. Wudy, Eva Jüttner, Joern Pons-Kühnemann, Christian Vokuhl, Michael C. Frühwald, Christoph Röcken, Lisa Schweizer, Pascal D. Johann, Antje Redlich, Matthias Mann, Michaela Kuhlen

## Abstract

Pediatric adrenocortical tumors (pACT) are biologically heterogeneous and incompletely classified by histopathology. To elucidate proteome-level tumor organization, we performed mass spectrometry–based proteomic profiling of 83 pACT and 43 normal adrenal samples. Unsupervised clustering identified four distinct molecular subtypes partially overlapping with histological diagnoses. These subtypes reflected discrete biological states: stromal–immune enrichment with reduced steroidogenesis, mitochondrial and steroidogenic activation, IGF/mTOR-driven anabolic reprogramming, and highly proliferative chromatin-remodeled carcinoma states. Proteomic clusters correlated strongly with endocrine phenotype, proliferative index, vascular invasion, and survival, outperforming conventional pathology.

A five-protein classifier (DAAM2, CIP2A, TSC2, PALS2, P3H1) reproduced subtype structure with 92.8% cross-validated accuracy. Cluster-specific analysis therapeutic vulnerabilities, including IGF-axis activation and epigenetic and DNA replication targets in aggressive tumors. These data establish a proteome-based classification of pACT integrating metabolic, proliferative, immune features, providing a framework for molecularly guided risk assessment and precision therapy in this rare cancer.

**Statement of significance:** Proteome-based classification of pediatric adrenocortical tumors identifies four functional tumor states that integrate metabolism, proliferation, and lineage identity with clinical outcomes. A minimal protein classifier and subtype-specific therapeutic vulnerabilities define a translational framework for molecular stratification and precision therapy in this rare endocrine malignancy.

## Introduction

Adrenocortical tumors (ACT) in children and adolescents are exceedingly rare, yet represent one of the most challenging pediatric endocrine malignancies. Their incidence is estimated at ∼0.2–0.3 cases per million children annually worldwide. There is a striking geographic clustering in southern Brazil due to a founder *TP53* variant (p.R337H)^1,2^. Pediatric ACT (pACT) encompass a clinical spectrum ranging from benign adenomas (ACA) to aggressive adrenocortical carcinomas (ACC). More than 80% are hormonally active, typically presenting with virilization, Cushing syndrome, or combined androgen–cortisol excess, which not only leads to significant morbidity but can also accelerate tumor detection^1,3–5^.

Despite decades of research, accurate diagnosis and risk stratification remain major hurdles in pACT management. Histopathological classification, traditionally adapted from adult scoring systems such as Weiss and Van Slooten, has limited reliability in children and adolescents. The pediatric-specific Wieneke criteria improved discrimination between ACA and ACC, but prognostic accuracy remains suboptimal^6–8^. Clinical factors such as age, stage, and extent of resection strongly influence outcome, yet patients with similar clinicopathological profiles may follow divergent clinical courses^9,10^. Current systemic therapies for advanced disease rely on mitotane and cisplatin-based regimens largely extrapolated from adult ACC protocols, with dismal long-term survival in metastatic disease (5-year survival <20%)^1,11^. There is an urgent need for molecularly informed diagnostic and prognostic biomarkers, as well as novel therapeutic targets^12^.

Genomic and transcriptomic studies have substantially advanced our understanding of pACT biology^3,13–16^. Comprehensive sequencing revealed recurrent alterations in *TP53*, *ATRX*, and *CTNNB1*, frequent loss of heterozygosity at 11p, and nearly universal IGF2 overexpression^3^. These discoveries underscore the developmental and genetic context of pACT, including links to hereditary cancer syndromes and imprinting disorders. More recently, transcriptomic profiling has identified prognostic gene expression signatures and pathway dependencies, including a 15-gene hub signature associated with 5-year event-free survival^14^. In adult ACC, integrated multi-omic characterization has defined molecular subtypes with distinct outcomes^17^. Nevertheless, transcriptional profiles do not fully capture the regulatory complexity that dictates tumor phenotype. Post-transcriptional modifications, protein abundance, and signaling pathway activation remain incompletely resolved at the RNA level^18,19^.

Proteomic approaches provide complementary insight, capturing the functional layer of molecular biology most proximal to cellular phenotype^20,21^. In adult ACT, mass spectrometry-based studies have revealed proteomic alterations, including upregulation of metabolic enzymes, cytoskeletal proteins, and mitochondrial pathways, as well as candidate prognostic markers^22–25^. Recent studies identified the IGF2 protein (juxtanuclear Golgi pattern) in 80% of carcinomas versus 0% of adenomas, establishing it as the most discriminating diagnostic biomarker at the protein level^25^. Additionally, a mass spectrometry study identified 64 upregulated and 48 downregulated proteins distinguishing ACC from adenoma, with evidence of Warburg effect metabolic reprogramming^24^. Jang et al. further demonstrated that proteomic profiles predict survival in ACC, highlighting their potential for clinical translation^26^. In contrast, pACT have not been studied yet at the proteomic level.

Together, these gaps provide a compelling rationale for systematic proteome profiling in pACT. By integrating quantitative proteomics with clinicopathological data, such studies may (i) improve the discrimination between adenomas and carcinomas, (ii) refine prognostic stratification, and (iii) uncover therapeutic vulnerabilities.

Here, we report a comprehensive proteomic analysis of pACT, establishing the largest protein expression dataset to date in this rare tumor entity and providing novel insights into tumor biology, biomarkers, and therapeutic opportunities.

## Results

### Study cohort and proteome profiling of pediatric adrenocortical tumors and matched normal adrenal tissue

We analyzed a clinically annotated cohort of children and adolescents diagnosed between 1999 and 2019, comprising 83 pACT samples from 72 patients and 43 matched normal adrenal cortex specimens from the same patients (Figure 1A). Tumors were classified according to the Wieneke criteria by centralized reference pathology review into ACC (n = 39), ACA (n = 23), and ACX (n = 15). Median age at diagnosis was 3.5 years (range 0.3–17.8), with a female-to-male ratio of 2.3:1. The majority of tumors were hormonally functional (77%; 54/70 with available endocrine phenotype data), with virilization as the predominant phenotype (n = 34), followed by combined virilization and Cushing syndrome (n = 13), Cushing syndrome alone (n = 7), and non-functional tumors (n = 16). Clinical and pathological characteristics are summarized in Supplementary Table S1.

**Figure 1.**
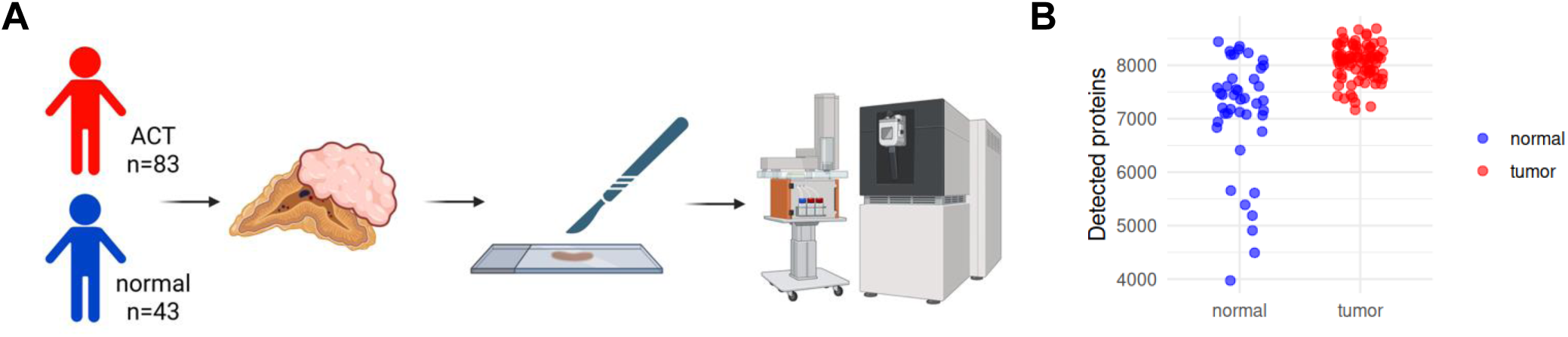
Cohort overview, sample processing workflow and proteome depth. (A) Schematic overview of cohort composition and proteomics workflow. Formalin-fixed, paraffin-embedded (FFPE) tissue from pediatric adrenocortical tumor specimens and matched non-neoplastic adrenal cortex were collected at surgery, followed by standardized macrodissection, protein extraction and digestion, and quantitative liquid chromatography – mass spectrometry-based proteome profiling. Partly created with BioRender. (B) Number of protein groups detected per sample, shown for normal adrenal cortex (blue), and tumor tissue (red). Each dot represents one sample. Protein counts are derived from the unfiltered raw expression matrix prior to any quality or completeness filtering.

Proteome profiling yielded a mean of 8,064 proteins per tumor sample (range 7,165–8,686) and 7,123 per normal adrenal cortex sample (range 3,973–8,442). In total, 10,714 non-redundant protein groups were quantified across the cohort. Tumor samples displayed significantly higher proteome depth than matched normal adrenal tissue (mean difference: 941 proteins; p < 0.001; Figure 1B, Supplementary Figure S1A), with greater inter-sample variability in normal tissue (SD 1,068 vs. 328), likely reflecting heterogeneity in cortical zone composition. Of 10,714 protein groups, 216 (2.0%) were removed due to insufficient detection across both sample groups. A strict completeness filter (≥50% of samples in each group) retained 7,035 proteins for all multivariate and visualization analyses. A parallel liberal filter retained an additional 3,463 group-specific proteins, predominantly tumor-exclusive, yielding 10,498 proteins available for differential abundance analysis (Supplementary Figure S1B, C). Remaining missing values in the liberally filtered matrix were imputed using a sample –wise Gaussian left-tail approach prior to statistical modeling. Log₂ transformation effectively corrected the strongly right-skewed raw intensity distributions (Supplementary Figure S1D), and per-sample log₂ intensity distributions were highly consistent across all analyzed samples (Supplementary Figure S1E).

### Tumor and normal adrenal tissues show distinct global proteome structure and broad differential protein abundance

We first examined whether global protein abundance profiles distinguish tumors from normal adrenal cortex. Unsupervised visualization and clustering of all samples revealed a clear separation of tumor and normal tissues. UMAP projection showed that the dominant axes of proteomic variation align with the tumor identity (Figure 2A), and sample-to-sample distance profiling together with hierarchical clustering identified two major branches corresponding to tumor and normal samples (Figure 2B). Clustering of the 500 most variable proteins further emphasized coherent expression programs separating tumors from normal adrenal cortex (Supplementary Figure S2A).

**Figure 2.**
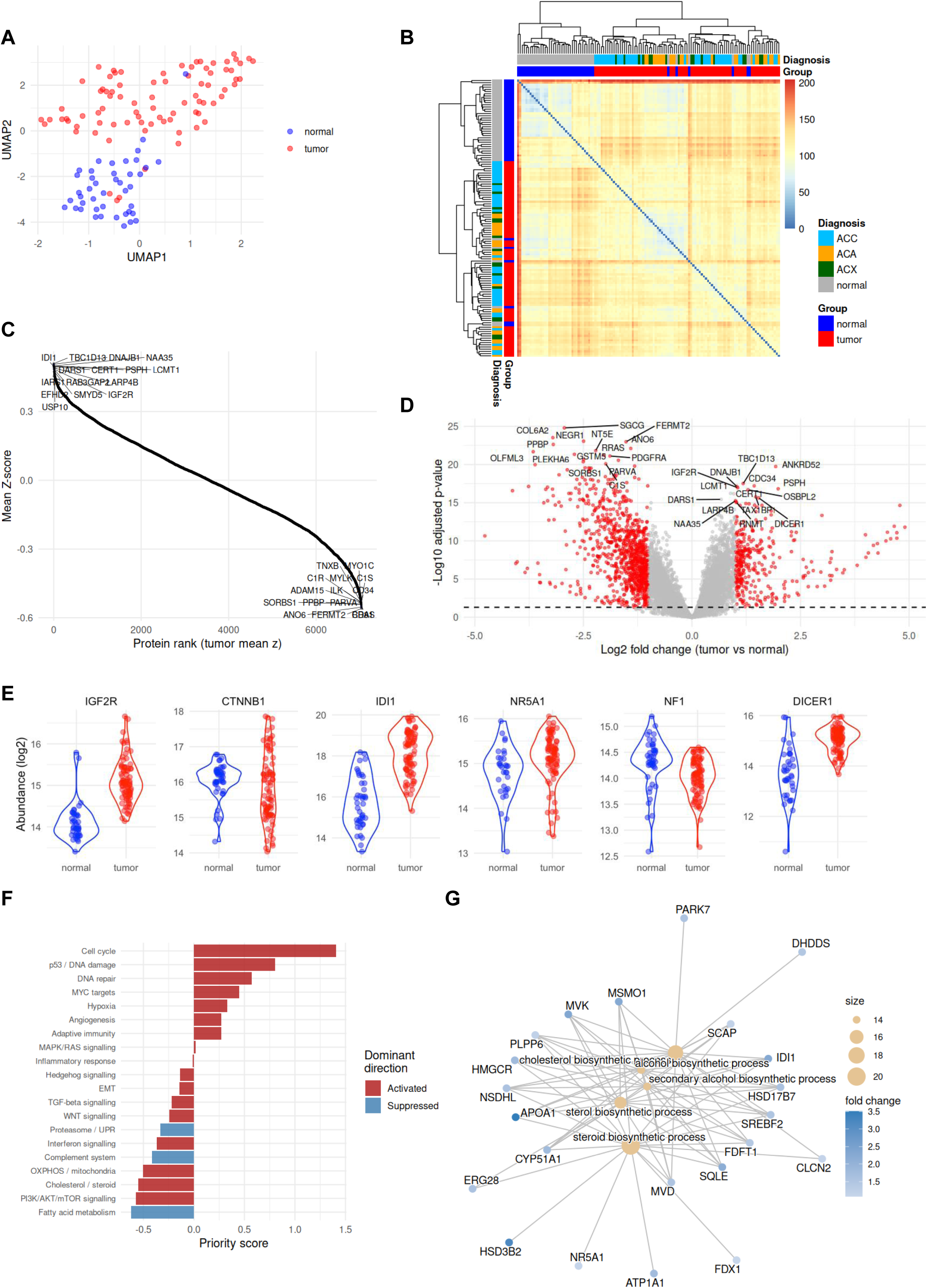
Global proteome structure, differential abundance, and functional enrichment in tumors versus normal adrenal tissue. Unless otherwise noted, visualization panels (A–C, E) use the strictly filtered matrix (7,035 proteins; ≥50% detection in each group); differential abundance and enrichment panels (D, F, G) use the liberally filtered, imputed matrix (10,498 proteins; ≥3 valid values in at least one group). (A) Uniform Manifold Approximation and Projection (UMAP) of the analysis-only dataset (tumor and normal samples; empty controls excluded). UMAP was computed on the Z-score-normalized protein abundance matrix. Each point represents one sample; color denotes group (tumor vs. normal). (B) Sample-to-sample Euclidean distance heatmap with hierarchical clustering. Annotation bars indicate group (tumor/normal) and diagnosis category (ACC/ACA/ACX/normal). (C) Tumor-centered protein ranking plot based on mean tumor Z-scores. For each protein, abundances were Z-score normalized across samples and averaged across tumor samples; proteins were ranked by this mean. Selected proteins are labeled. (D) Volcano plot of differential protein abundance between tumors and normal adrenal cortex (limma; liberally filtered, imputed matrix). The x-axis shows log2 fold change (tumor vs. normal) and the y-axis −log10 adjusted p-value (Benjamini–Hochberg); points are colored by significance (FDR < 0.05, |log2 FC| ≥ 1). (E) Log2-transformed abundance for selected proteins from the tumor–normal comparison: IGF2R, CTNNB1, IDI1, NR5A1, NF1, and DICER1. Violin shapes depict the distribution; individual points represent samples. Observed values only (no imputation). (F) Multi-source convergence priority ranking of biological themes in tumor versus normal adrenocortical tissue. Horizontal bar plot showing the heuristic composite priority score for each a priori-defined biological theme, ranked in descending order. Bar color indicates the dominant direction of regulation across significant pathways assigned to that theme (red, Activated; blue, Suppressed, based on mean normalized enrichment score, NES). The priority score is a dimensionless composite z-score integrating cross-database convergence (number of independent enrichment sources with at least one significant pathway for the theme) and effect magnitude (mean absolute NES across significant GSEA pathways assigned to the theme). (G) GO Biological Process category–protein association network (cnetplot) for proteins with increased abundance in tumors. Term nodes represent enriched GO BP categories; protein nodes represent contributing members; color encodes log2 fold change. The network highlights the interconnected enzymes of the mevalonate–cholesterol–steroid biosynthetic axis.

To identify proteins consistently altered in tumors, we ranked all proteins by their mean tumor-centered Z-score. This analysis highlighted a set of proteins uniformly elevated across tumors, including TBC1D13, DNAJB1, LCMT1, CERT1, IGF2R, and IDI1, and a corresponding group consistently reduced, including FERMT2, SORBS1, RRAS, OLFML3, and TNXB (Figure 2C), providing reference candidates for subsequent analyses.

Differential abundance analysis using a linear model with empirical Bayes moderation quantified the extent of tumor-normal divergence. At FDR < 0.05 and |log₂FC| ≥ 1, 342 proteins were significantly increased and 956 significantly decreased in tumors, yielding 1,298 differentially abundant proteins in total. The volcano plot demonstrated widespread dysregulation with strong statistical support (Figure 2D). The most significantly increased proteins included ANKRD52, TBC1D13, CDC34, IGF2R, and DNAJB1 (adjusted p < 10⁻¹⁷), whereas the most significantly decreased proteins, SGCG, COL6A2, NEGR1, FERMT2, and OLFML3 (adjusted p < 10⁻²²), were predominantly linked to extracellular matrix (ECM) organization and adhesion programs. Representative examples in part linked to pACT pathogenesis illustrating the magnitude and consistency of protein differences are shown in Figure 2E.

Pathway-level analyses condensed the protein-level alterations into coherent biological themes, revealing a fundamental metabolic and signal transduction rewiring in pACTs. An integrated pathway priority analysis identified p53/DNA damage, DNA repair, PI3K/AKT/mTOR signaling, TGF-β signaling, WNT signaling, MYC targets, cell cycle, cholesterol/steroid metabolism, and OXPHOS/mitochondria and hypoxia programs as coordinately activated, while proteasomal programs, fatty acid metabolism, and complement system were suppressed (Figure 2F; Supplementary Methods).

Central to this reprogramming was a pronounced activation of the mevalonate–cholesterol–steroid biosynthetic axis. GO biological process ORA of proteins increased in tumors revealed over-representation of small molecule and steroid biosynthetic processes, alcohol and sterol metabolic processes, cholesterol metabolic and biosynthetic processes, response to reactive oxygen species, DNA replication initiation, and platelet aggregation (Supplementary Figure S2B-E). Reactome ORA further confirmed activation of steroid metabolism, cholesterol biosynthesis, SREBP/SREBF-regulated cholesterol biosynthesis and gene expression, lanosterol biosynthesis, ATR activation in response to replication stress, ECM reorganization and integrin cell surface interactions (Supplementary Figure S2D). Network visualization demonstrated that these cholesterol– and steroid-related processes were driven by a highly interconnected enzyme core, including enzymes of the mevalonate pathway (HMGCR, MVK, MVD, IDI1, FDFT1, SQLE, PLPP6), post squalene cholesterol biosynthesis (CYP51A1, MSMO1, NSDHL, HSD17B7, ERG28), the (cholesterol sensor/transcription factor complex (SREBF2, SCAP), steroidogenesis (NR5A1, HSD3B2, FDX1), and cholesterol transport (APOA1), collectively indicating coordinated activation of the mevalonate–cholesterol–steroid axis (Figure 2G).

This metabolic shift was accompanied by parallel rewiring of proliferative and stress-response signaling. GSEA using MSigDB Hallmark gene sets confirmed activation of MYC targets, mTORC1 signaling, cholesterol homeostasis, unfolded protein response, and E2F targets, alongside suppression of epithelial–mesenchymal transition, apical junction, complement, inflammatory response, and angiogenesis pathways in tumors relative to normal adrenal tissue (Supplementary Figure S2B).

Collectively, these analyses define pACTs as tumors characterized by a coupled metabolic and signal transduction reprogramming: coordinated activation of the mevalonate–cholesterol–steroid biosynthetic network is coupled with engagement of MYC-, mTORC1-, and cell cycle–driven proliferative programs, while ECM organization, stromal adhesion, immune surveillance, and developmental signaling programs are concurrently dismantled.

### Proteomic heterogeneity across tumor subgroups and adrenal zonation features

Having established robust global differences between tumors and normal adrenal cortex, we next examined whether proteomic variation also stratifies histological tumor subgroups. Tumor-only samples were compared across ACC, ACA, and ACX using a linear model with empirical Bayes variance moderation (overall F-test). The top 50 discriminating proteins revealed partially overlapping abundance patterns across ACC, ACA and ACX, with incomplete separation between diagnostic categories (Figure 3A). Many top-ranked proteins mapped to proteostasis, mitochondrial, and biosynthetic pathways, suggesting differences in cellular state across diagnostic categories despite substantial inter-group overlap.

**Figure 3.**
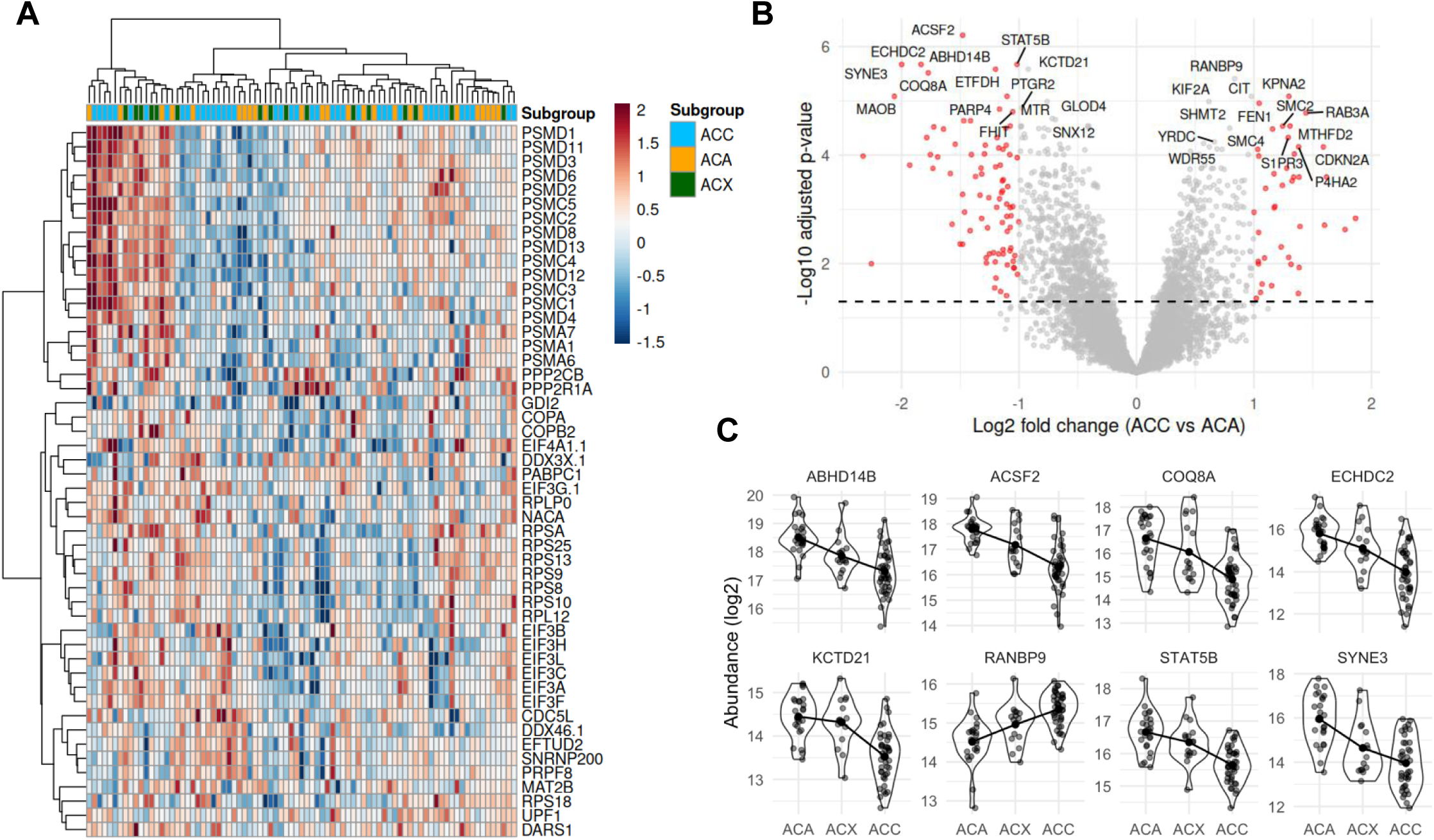
Proteomic heterogeneity across histopathological subgroups (tumor-only analysis). All panels use the strictly filtered, log2-transformed tumor-only matrix (7,035 proteins; ≥50% detection in each group), without imputation. (A) Heatmap of the top 50 proteins discriminating tumor subgroups (F-test across ACC/ACA/ACX, BH-adjusted). Proteins were selected by F-statistic and displayed as Z-score-normalized values; hierarchical clustering of samples and proteins used Ward’s method. Annotation bar indicates diagnosis (ACC blue, ACA orange, ACX green). (B) Volcano plot for the pairwise ACC vs. ACA contrast (limma with empirical Bayes moderation; tumor-only matrix). The x-axis shows log2 fold change (ACC vs. ACA) and the y-axis −log10 adjusted p-value (BH). Points are colored by significance (FDR < 0.05, |log2 FC| ≥ 1); the 15 most significant proteins in each direction are labeled. (C) Violin/jitter plots of the top 8 proteins with monotonic abundance trends across the ordered ACA → ACX → ACC sequence (ordinal limma model, BH-adjusted). Each panel shows log₂ abundance per subgroup; the overlaid line connects subgroup means. Proteins shown: ABHD14B, ACSF2, COQ8A, ECHDC2, KCTD21, RANBP9, STAT5B, SYNE3.

A focused pairwise contrast between ACC and ACA delineated a carcinoma-specific proteomic signature (Figure 3B). ACC tumors exhibited coordinated increase in proteins involved in oxidative phosphorylation, ATP synthesis, ribosome biogenesis, and RNA processing. Gene set enrichment analysis (GSEA) using ranked moderated t-statistics confirmed enrichment of mitochondrial respiration and translational programs in ACC relative to ACA and reinforced activation of biosynthetic and energy-generating pathways (Supplementary Figure S3A, B). These findings indicate that carcinomas are characterized by enhanced metabolic and proliferative capacity compared with adenomas.

We next explored whether protein abundance changes follow a graded trajectory across the histological sequence ACA–ACX–ACC. Ordinal trend modeling identified proteins with monotonic increases or decreases, including ABHD14B, ACSF2, COQ8A, ECHDC2, KCTD21, RANBP9, STAT5B, and SYNE3 (Figure 3C). To assess whether these trends reflect a global gradient, we tested the correlation between the first ten principal components and ordinal subgroup coding (Spearman, BH-adjusted). PC1 (10.5% variance) showed no significant association (rho = 0.19, adj. p = 0.29; Supplementary Figure S3C–D), confirming that the dominant proteomic variance axis is not aligned with the ACA–ACX–ACC sequence. In contrast, PC2 (9.1% variance; rho = 0.64, adj. p < 0.0001) and PC5 (3.9%; rho = 0.40, adj. p = 0.00086) captured significant monotonic gradients. Proteins driving the PC2 gradient toward ACC were enriched for DNA replication, repair, and chromatin remodeling (e.g. DNMT1, FEN1, SMC2/4, MCM2–6), while those decreasing toward ACC were dominated by mitochondrial and organic acid metabolism (e.g. TK2, MCEE, LACTB, CYP27A1; Supplementary Figure S3E–F). Thus, ACA-to-ACC progression is associated with a gain of proliferative and genome-maintenance capacity coupled with a loss of differentiated metabolic programs, but this gradient is embedded in secondary variance components rather than dominating the overall proteomic landscape.

Finally, we examined whether inter-tumor heterogeneity relates to adrenal cortical lineage features. Zonation marker scores for zona glomerulosa (ZG), zona fasciculata (ZF), and zona reticularis (ZR) were calculated as the mean Z-score of the best-detected markers per zone (Supplementary Table S2). Unsupervised clustering of these markers demonstrated clear separation of zonal blocks (Supplementary Figure S4A), with heterogeneous but structured patterns across tumors. Proportional zonation compositions derived via softmax transformation revealed mixed zone signatures in most tumors (Supplementary Figure S4B), while ternary plots illustrated subgroup-associated shifts in zonal representation, with ACC tumors exhibiting comparatively constrained patterns (Supplementary Figure S4C). These observations suggest partial retention and/or reconfiguration of adrenal cortical lineage identity within pACT. Collectively, while histological categories capture discrete proteomic differences, the absence of a dominant global gradient across the tumor spectrum suggests additional layers of tumor-intrinsic heterogeneity beyond morphology.

### Unsupervised proteome-based clustering defines four molecular tumor subtypes with distinct functional programs and lineage features

To resolve tumor-intrinsic heterogeneity independent of histopathological diagnosis, we applied consensus clustering (ConsensusClusterPlus, Ward’s D2, Euclidean distances, 1,000 resamplings) to the tumor-only proteome. The consensus cumulative distribution function (CDF) and delta area plots identified a four-cluster solution as the most stable partition (Supplementary Figure S5A-C), yielding clusters C1–C4 comprising 25, 14, 27, and 17 tumors, respectively. The mean silhouette width of 0.84 indicated robust separation between clusters. In UMAP space, these proteome-defined clusters segregated clearly, while diagnostic labels were distributed non-uniformly across clusters (Figure 4A), demonstrating that cluster structure does not recapitulate histopathology.

**Figure 4.**
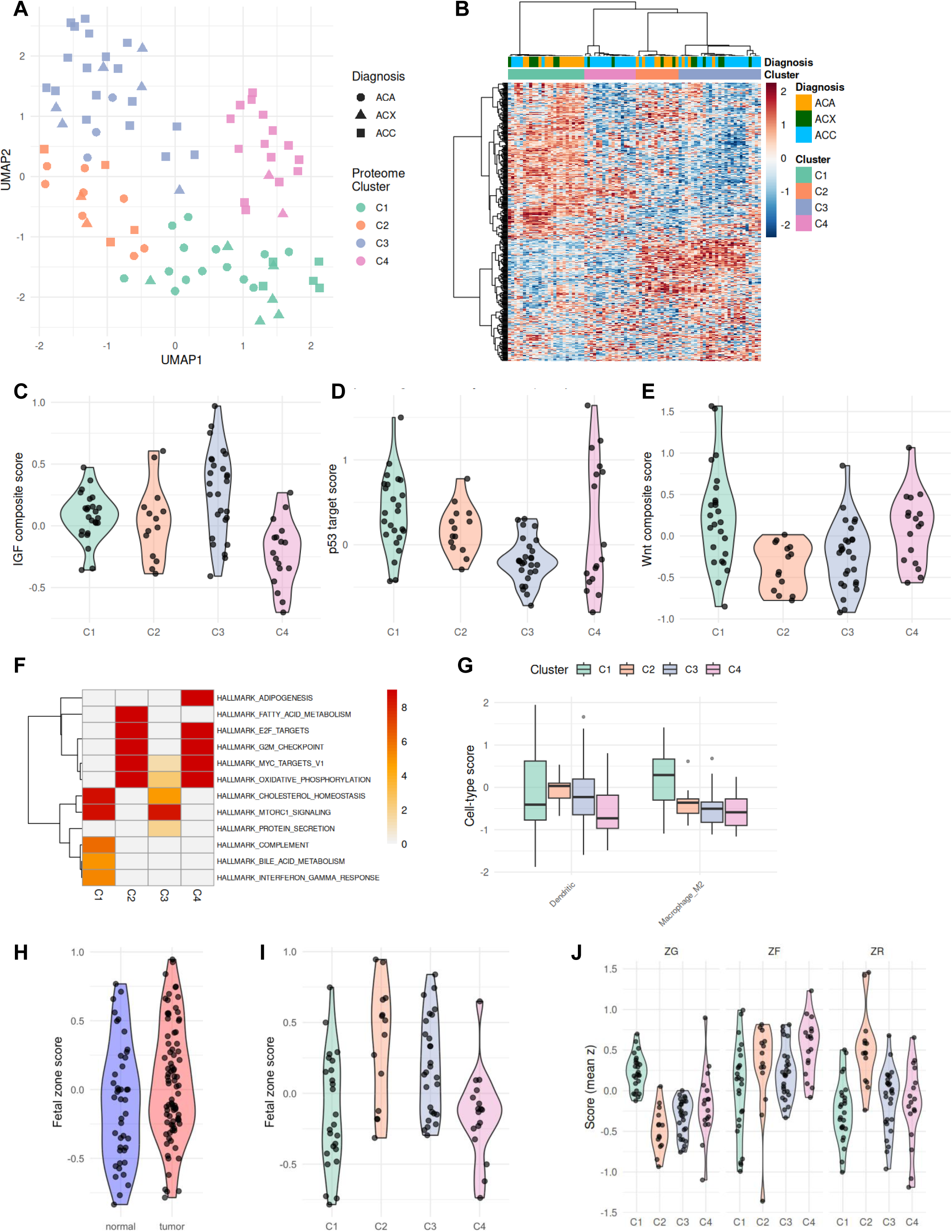
Proteome cluster-specific functional programs, oncogenic pathway scores, immune microenvironment, and adrenal zonation features. All panels use the strictly filtered, log2-transformed tumor-only matrix (7,035 proteins), without imputation. Consensus clustering was performed on the top MAD-ranked proteins (ConsensusClusterPlus, Ward’s D2, Euclidean distances, 1,000 resamplings, Pearson distance); k = 4 was selected based on consensus CDF and delta area criteria (Supplementary Figure S6). (A) UMAP of the tumor-only proteome colored by proteome cluster (C1–C4); diagnosis is encoded by point shape (circle = ACA, triangle = ACX, square = ACC). (B) Heatmap of the most variable proteins across all tumor samples, displayed as Z-score-normalized values. Hierarchical clustering of samples used Ward’s method. Annotation bars indicate proteome cluster (C1–C4) and diagnosis (ACC/ACA/ACX). (C) Composite IGF signaling proteomic score per sample, shown as violin plots stratified by cluster. Scores were computed as the mean Z-score of curated pathway member proteins detected in the dataset. (D) Composite p53 target proteomic score per sample, stratified by cluster. Computed as in panel C. (E) Composite Wnt pathway proteomic score per sample, stratified by cluster. Computed as in panel C. (F) MSigDB Hallmark gene set enrichment heatmap across clusters. Enrichment was performed per cluster (cluster vs. rest, limma; BH-adjusted). Color encodes normalized enrichment direction and significance; rows represent Hallmark gene sets, columns represent clusters. (G) Immune cell type marker scores per cluster, shown as boxplots. Only dendritic cell and M2 macrophage signatures were detected at sufficient marker coverage (≥2 markers per cell type) for reliable scoring. (H) Fetal adrenal cortex proteomic scores per sample, shown as violin plots stratified by tumor versus normal and by cluster. Scores were computed as the mean Z-score of detected marker proteins. (I) Ternary plot of sample-wise adrenal zone compositions (pZG, pZF, pZR) stratified by proteome cluster. Each vertex represents 100% contribution of one zone; scores were converted to proportions via softmax transformation. (J) Individual ZG, ZF, and ZR zone scores per sample shown as violin plots stratified by cluster, illustrating the cluster-specific lineage biases underlying the ternary compositions in panel I.

Cluster composition revealed partial but incomplete overlap with histopathological diagnosis. C1 and C2 were enriched for ACA (56% and 57%, respectively), C3 was predominantly ACC (67%), and C4 comprised almost exclusively ACC tumors (88%), with only two ACX cases and no ACA. Structured expression programs underlying these groups were evident in the heatmap of the most variable proteins (Figure 4B), with expanded marker panels and enrichment analyses provided in Supplementary Figure S6.

Pathway-level interrogation revealed distinct functional states across clusters. Composite scores of key pACT pathways quantified these distinctions (Figure 4C–E). IGF signaling activity differed significantly across clusters (KW p < 0.0001), peaking in C3 and being lowest in C4. p53 target scores (KW p = 0.0003) were highest in C1 (and lowest in C3, while Wnt pathway scores (KW p = 0.0004) were elevated in C1 and lowest in C2. Furthermore, C1 was characterized by enrichment of phagocytosis and actin cytoskeleton remodeling programs alongside relative suppression of sterol biosynthesis and mTORC1 signaling. C2 displayed strong activation of mitochondrial organization, oxidative phosphorylation, and respiratory chain programs, consistent with enhanced bioenergetic capacity. C3 was distinguished by elevated levels of cholesterol and sterol biosynthesis proteins and mTORC1 activation, consistent with anabolic steroidogenic programs. C4 exhibited the most proliferative phenotype, with enrichment of E2F targets, G2M checkpoint, MYC targets, and chromatin organization pathways, accompanied by relative suppression of TCA cycle activity, consistent with a highly dedifferentiated carcinoma state (Figure 4F; Supplementary Figure S6A-E). Immune-related features were comparatively limited. Among evaluated immune cell signatures, only M2 macrophage scores varied significantly (KW p < 0.0001), being highest in C1 and progressively decreasing across C2, C3, and C4 (Figure 4G). Dendritic cell signatures did not differ significantly (KW p = 0.36). The enrichment of M2-associated markers in C1 aligns with its phagocytosis and cytoskeletal remodeling profile and likely reflects differences in stromal composition rather than classical tumor-intrinsic immune activation.

Of the 7 fetal adrenal panel proteins, 4 were quantified (IGF2, CYP17A1, CYB5A, SULT2A1). Fetal adrenal scores showed a trend toward higher values in tumors compared with normal adrenal tissue (Wilcoxon p = 0.066; Figure 4H), suggesting that this overall difference may be driven by a subset of tumors. Indeed, when stratified by proteome cluster, fetal scores differed significantly (KW p = 0.00055; Figure 4I), with clusters C2 and C3 showing the highest values. A mature cortex score could not be computed as most panel members (including CYP11B2, MC2R, and MRAP) fell below the detection limit of the label-free workflow.

Finally, adrenal zonation marker scoring revealed distinct lineage biases (Figure 4I, J; Supplementary Figure S7). C1 exhibited a ZG-like predominance, C2 showed a ZR-like identity (lowest ZG representation), and C3 and C4 were ZF-dominant, with C4 demonstrating the most pronounced fasciculata-like signature. These lineage patterns suggest that proteome-defined tumor subtypes integrate functional metabolic programs with retained or reconfigured adrenal cortical lineage identity.

Collectively, these analyses define four proteome-based tumor subtypes integrating metabolic specialization, proliferative capacity, immune context, and adrenal lineage features, extending histopathologic classification into a multidimensional molecular framework.

### Proteome-based clusters reflect distinct clinical characteristics

Having defined four proteome-based tumor subtypes, we next examined their clinical correlates. Age at diagnosis differed markedly across clusters (Kruskal-Wallis p < 0.0001; Figure 5A): C1 and C3 were strongly enriched for young children aged 0–3 years (71% and 58%, respectively), whereas C2 and C4 comprised predominantly older patients (>4 years; 62% and 64%, respectively). In contrast, sex distribution did not differ significantly (Fisher’s exact p = 0.598; Figure 5B), with females representing the majority in all clusters (62–85%).

**Figure 5.**
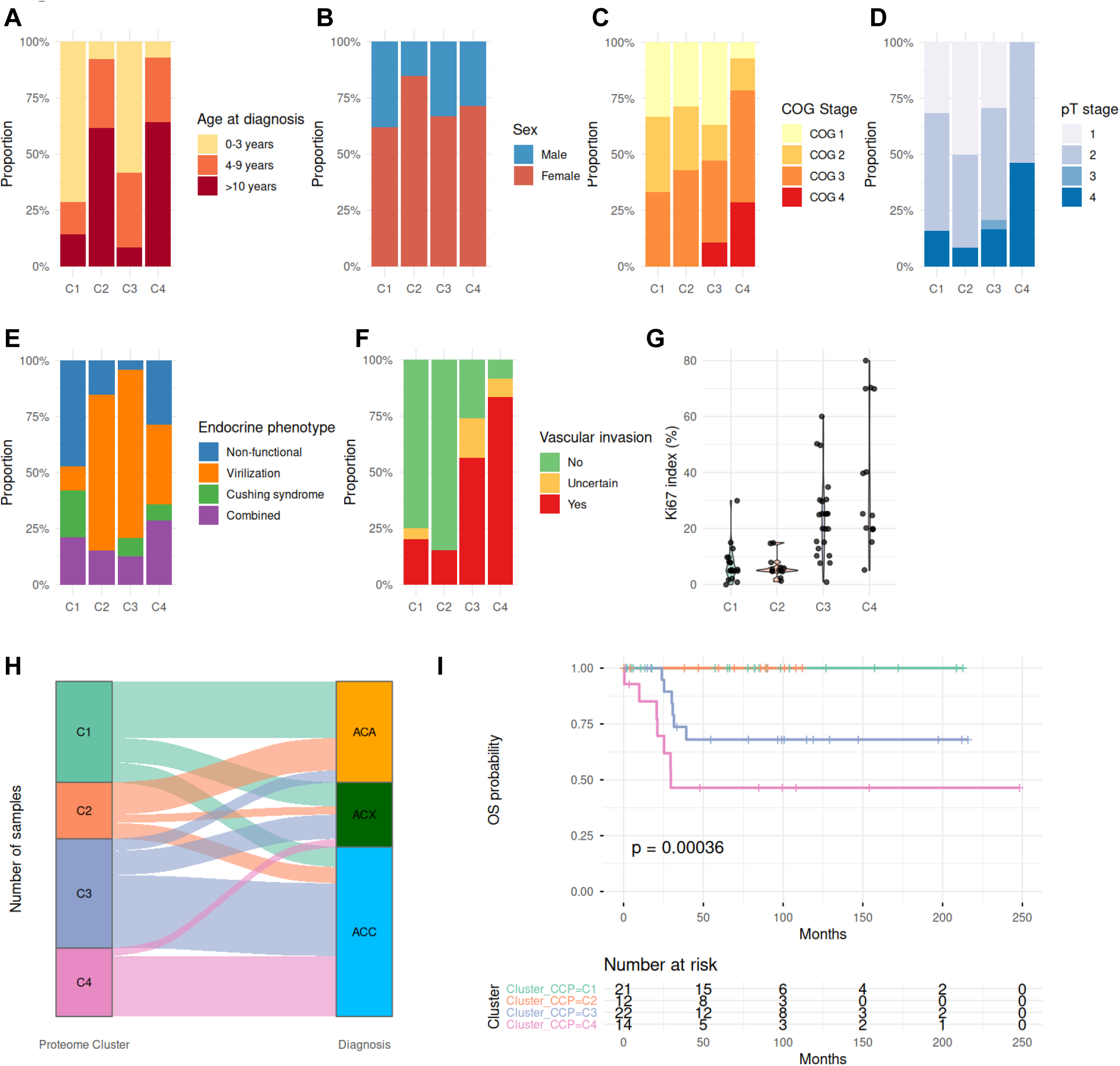
Clinical and pathological correlates of proteome-based cluster membership. Clinical variables were assessed across the four proteome clusters (C1–C4) using Fisher’s exact test for categorical variables and Kruskal-Wallis test for continuous variables. (A) Age at diagnosis stratified by cluster, categorized as 0–3 years, 4–9 years, and >10 years (Fisher’s exact p < 0.0001). Stacked proportion bars; same format for panels B–F. (B) Sex distribution across clusters (Fisher’s exact p = 0.598). (C) COG stage distribution across clusters (Fisher’s exact p = 0.410). (D) pT stage distribution across clusters (Fisher’s exact p = 0.092). (E) Endocrine phenotype distribution across clusters, categorized as non-functional, virilization, Cushing syndrome, and combined (Fisher’s exact p = 0.0009). (F) Vascular invasion status across clusters (Fisher’s exact p < 0.0001). (G) Ki67 proliferation index (%) per cluster shown as violin plots with individual data points. Median Ki67: C1 = 5%, C2 = 5%, C3 = 25%, C4 = 25% (mean C4 = 37%). Kruskal-Wallis p < 0.0001. (H) Sankey diagram illustrating the mapping of samples from proteome cluster (C1–C4) to histological diagnosis (ACA, ACX, ACC). (I) Kaplan–Meier overall survival curves stratified by proteome cluster. The x-axis shows time in months (0–250); the y-axis shows OS probability (log-rank p = 0.00036). Numbers at risk are shown below the plot.

Tumor stage parameters showed more nuanced patterns. Neither COG stage nor pT stage reached statistical significance across clusters (Fisher’s exact p = 0.410 and p = 0.092; Figure 5C, D), although C4 displayed a trend toward higher-stage disease (50% COG stage 3–4; 43% pT4). In contrast, endocrine phenotype differed significantly (Fisher’s exact p = 0.0009; Figure 5E). C2 and C3 were enriched for Cushing syndrome (69% and 63% of evaluable cases), whereas C1 was predominantly non-functional (68%), consistent with its relative suppression of steroid biosynthetic programs.

Markers of aggressive tumor behavior were strongly cluster-associated. Vascular invasion varied significantly across clusters (Fisher’s exact p < 0.0001; Figure 5F), being largely absent in C1 and C2 (75% and 85% without vascular invasion) but frequent in C3 and C4 (54% and 71%). Ki67 proliferation index and necrosis both differed robustly (Kruskal-Wallis p < 0.0001 and Fisher’s exact p < 0.0001; Figure 5G, Supplementary Figure S8A). C1 and C2 showed low proliferative activity (median Ki67 5%) and low rates of necrosis (32% and 18%), C3 displayed intermediate-to-high proliferation (median Ki67 25%) with necrosis in 71% of cases, and C4 exhibited the highest proliferative indices (median 25%, mean 37%) with near-universal necrosis (93%, Supplementary Figure S8A), consistent with its highly proliferative molecular profile.

Cluster membership was significantly associated with histological diagnosis (Fisher’s exact p < 0.0001; Figure 5H), yet the distribution confirmed only partial concordance between morphology and proteome-defined subtypes. Importantly, overall survival differed significantly across clusters (log-rank p = 0.00036; Figure 5I), with C1 and C2 showing the most favorable outcomes and C4 the poorest prognosis. Notably, survival stratification by histological diagnosis alone yielded weaker separation (log-rank p = 0.0023; Supplementary Figure S8B), indicating that proteome-based clustering provides superior prognostic resolution compared with histopathology alone.

Together, these findings demonstrate that proteome-defined tumor subtypes integrate molecular programs with demographic, endocrine, and pathological features, translating into distinct clinical trajectories.

### Clinical translatability of proteome-based tumor subtypes: a compact protein classifier and cluster-specific therapeutic vulnerabilities

To evaluate whether proteome-defined tumor subtypes can be approximated by a clinically tractable marker set, we derived a compact classifier using greedy forward feature selection with leave-one-out cross validation (LOO-CV) and a nearest-centroid model. Candidate features were drawn from the top 50 one-vs-rest differentially abundant proteins per cluster (FDR < 0.05, log₂FC > 0.5), yielding up to 200 non-redundant candidates. The optimal panel size was defined as the smallest set achieving ≥95% of maximal cross-validated accuracy.

A five-protein panel comprising DAAM2, CIP2A, TSC2, PALS2, and P3H1 achieved a LOO-CV accuracy of 92.8% (77/83 correctly classified tumors) (Figure 6A). Per-cluster sensitivities were 100%, 93%, 85%, and 94%, with specificities of 91%, 100%, 100%, and 99% for C1–C4, respectively. Classification relies on the combinatorial abundance pattern across all five proteins rather than on individual cluster-exclusive markers: DAAM2 is preferentially abundant in C1, CIP2A and TSC2 in C2, PALS2 in C2 and C3, and P3H1 in C3 and C4 (Supplementary Figure S9A–E). Notably, no single protein distinguished C4; instead, C4 tumors were classified by low abundance of DAAM2, CIP2A, and TSC2 together with high P3H1, a pattern consistent with the dedifferentiated molecular profile of this cluster. Permutation testing (1,000 permutations; null mean accuracy 25.1%) confirmed that classification performance significantly exceeded chance (p < 0.001; Supplementary Figure S9F). Hierarchical clustering and UMAP based solely on the five-protein matrix recapitulated full-proteome cluster structure (Figure 6B, C), demonstrating that a minimal marker set preserves the major biological stratification.

**Figure 6.**
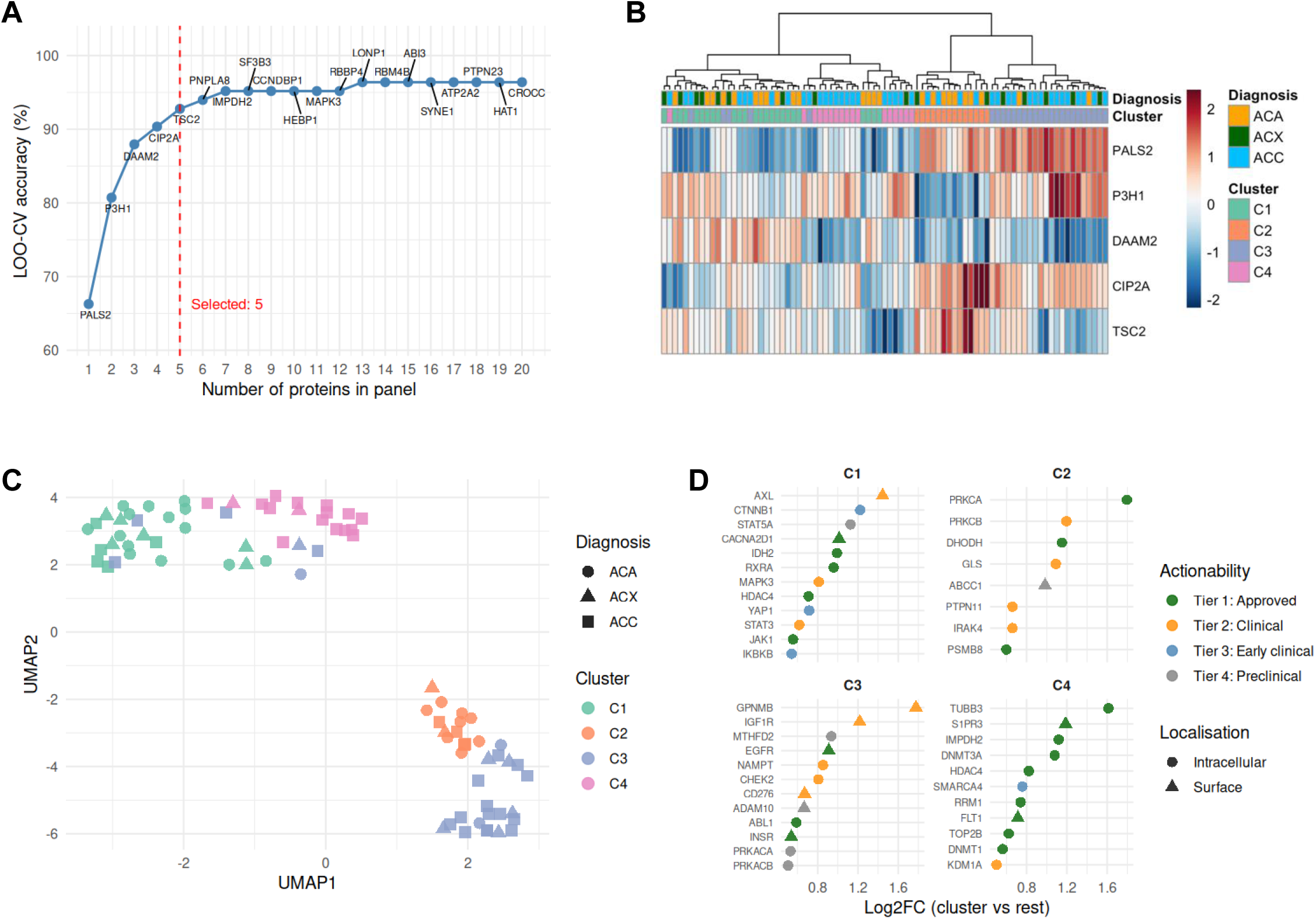
Compact protein classifier and cluster-specific therapeutic target landscape. (A) LOO-CV classification accuracy (%) as a function of panel size during greedy forward feature selection with a nearest-centroid classifier. Candidates comprised the top 50 differentially abundant proteins per cluster (one-vs.-rest; FDR < 0.05, log2 FC > 0.5), de-duplicated across clusters. Each point is labeled with the added protein; the red dashed line marks the selected five-protein panel (LOO-CV accuracy = 92.8%). (B) Heatmap of Z-score-normalized abundance of the five panel proteins (DAAM2, CIP2A, TSC2, PALS2, P3H1) across all tumor samples, hierarchically clustered (Ward’s method) with cluster and diagnosis annotation bars. (C) UMAP of tumor samples computed exclusively from the five-protein panel. Colors indicate proteome cluster (C1–C4); shape indicates diagnosis (circle = ACA, triangle = ACX, square = ACC). (D) Cluster-specific therapeutic target landscape: dot plot of log2 fold change (cluster vs. rest) for actionable proteins in each cluster. Dot color indicates actionability tier (Tier 1: approved; Tier 2: clinical trials; Tier 3: early clinical; Tier 4: preclinical); dot shape indicates localization (surface vs. intracellular).

Beyond classification, we systematically interrogated cluster-specific proteomes for potentially actionable therapeutic targets using a curated druggability framework stratified by target class, localization, and development stage (Figure 6D; Supplementary Figure S10). Distinct therapeutic vulnerabilities emerged across clusters.

C1 displayed the broadest actionable spectrum (24 candidates, including 13 Tier 1 targets), encompassing signaling mediators and transcriptional regulators such as MAPK3, AXL, CTNNB1, STAT3, YAP1, and HDAC4. C2 yielded 8 targets including PTPN11, PRKCA/B, DHODH, and IRAK4, consistent with its kinase-dependent metabolic phenotype. C3 identified 13 targets, including 6 surface-expressed and 2 antibody-drug conjugate-eligible proteins, notably IGF1R, NAMPT, ADAM10, and GPNMB. The enrichment of IGF1R is particularly relevant in the context of established IGF2 overexpression in pACT. C4 exhibited 11 targets with the highest Tier 1 proportion (9/11), dominated by epigenetic regulators (SMARCA4, HDAC4, DNMT3A, KDM1A) and DNA replication machinery components (TOP2B, RRM1), aligning with its chromatin remodeling and highly proliferative phenotype.

Collectively, these findings demonstrate that proteome-defined tumor subtypes are not only biologically distinct but can be approximated by a minimal diagnostic panel and linked to cluster-specific therapeutic vulnerabilities, providing a conceptual framework for molecularly guided stratification and hypothesis-driven therapeutic exploration in pACT.

## Discussion

ACT represent one of the biologically most heterogeneous endocrine malignancies. This is particularly true in pediatric patients where classic histopathological criteria incompletely capture malignant potential^1,9,10,12,27^. Over the past decade, genomic and transcriptomic studies have substantially advanced our understanding of ACT biology^3,13,14^. Integrative analyses in adult ACC, most prominently through TCGA, identified recurrent alterations in *TP53*, *CTNNB1*, *ZNRF3*, *CDKN2A*, and chromatin-remodeling genes, as well as transcriptome-based subgroups associated with proliferation and CpG island methylator phenotypes^17,28,29^. In pACT, genomic studies have emphasized germline and somatic *TP53* alterations, particularly the Brazilian founder variant, together with *IGF2* overexpression and methylation-based risk stratification^3,13,15,30^. More recently, RNA sequencing studies have proposed prognostic gene expression signatures and co-expression networks linked to steroidogenesis and cell cycle activity^14^.

Despite these advances, molecular stratification of ACT relies largely on upstream regulatory levels, DNA mutations, methylation, and RNA abundance, while the proteome, which is closest to the disease phenotype and represents the integrated functional output of these processes, has been comparatively understudied^18,19^. Prior proteomic investigations in adult ACC have predominantly focused on tumor-versus-normal comparisons, reporting dysregulation of steroid biosynthesis, lipid metabolism, and mitochondrial pathways^22–26^. However, these studies were limited by smaller cohort sizes and did not define reproducible tumor subtypes with integrated clinical and prognostic annotation. In pACT, no proteomic characterization has been reported to date.

Here, we demonstrate that whole-proteome profiling resolves four robust tumor subtypes that refine beyond histopathological classification and integrate metabolic specialization, proliferative signaling, immune context, and adrenal lineage identity. Importantly, these proteome-defined clusters were only partially concordant with morphological categories and provided more detailed prognostic stratification than histology alone. This observation suggests that functional molecular states captured at the protein level reflect clinically meaningful biology not fully resolved by current diagnostic frameworks^18–20^.

Rather than a simple linear malignancy continuum across ACA–ACX–ACC, our analyses revealed qualitatively distinct tumor subtypes. C2 and C3 were characterized by metabolically active steroidogenic programs, enriched for oxidative phosphorylation, mitochondrial organization, and cholesterol biosynthesis^31,32^. The anabolic and IGF-enriched phenotype of C3 aligns with established *IGF2* overexpression in pACT and may reflect sustained mTORC1-driven biosynthetic activity^3,33,34^. In contrast, C4 exhibited a highly proliferative, chromatin-remodeled state marked by *E2F* and *MYC* activation and the poorest survival outcomes. This profile complements prior genomic observations implicating chromatin regulators and cell cycle dysregulation in aggressive ACC^16,17,35^.

C1, enriched for actin remodeling, phagocytosis, and M2 macrophage-associated signatures, suggests a tumor subtype characterized by enhanced stromal interaction or altered microenvironment composition^36^. The enrichment of immune and cytoskeletal programs in this cluster underscores that proteomic profiling captures not only tumor cell–intrinsic programs but also tumor–microenvironment interplay, an aspect incompletely resolved by transcriptomics alone^37^.

These findings suggest that pACT biology is organized around discrete functional states rather than a uniform progression model. Proteomics thus provides a systems-level perspective that integrates metabolic, proliferative, and lineage programs into coherent tumor phenotypes. Our proteome-defined clusters do not simply recapitulate known genomic or methylation subgroups but instead functionally contextualize them. For example, the IGF1R enrichment observed in C3 corresponds with established *IGF2* overexpression, yet proteomics further reveals downstream anabolic cholesterol biosynthesis and mTORC1 activation as coordinated outputs. Similarly, the chromatin regulator enrichment in C4 complements genomic reports of alterations in epigenetic modifiers, suggesting that proteomic stratification captures the functional consequences of these alterations.

Importantly, while transcriptomic studies frequently stratify ACT by proliferation signatures, our data indicate that proliferative state represents only one dimension of tumor heterogeneity. Metabolic specialization, steroidogenic capacity, and lineage bias constitute additional independent organizing dimensions that are more directly observable at the protein level. Proteome-defined clusters were strongly associated with clinical parameters, including vascular invasion, necrosis, proliferative index, endocrine phenotype, and overall survival. Survival stratification by cluster exceeded that achieved by histological diagnosis alone, highlighting the potential of proteome-based classification to refine risk assessment.

The derivation of a five-protein classifier achieving 92.8% cross-validated accuracy demonstrates that proteomic stratification can be operationalized into a minimal marker panel compatible with immunohistochemical workflows in routine clinical practice. While prospective external validation remains necessary, this approach establishes a translational bridge from discovery proteomics to routine pathology.

Equally important is the identification of cluster-specific therapeutic vulnerabilities. The enrichment of IGF1R in C3 reinforces the rationale for IGF-axis targeting, whereas the concentration of epigenetic regulators and replication machinery components in C4 suggests potential sensitivity to chromatin-modifying agents or replication stress–targeting strategies^9,34,38^. Such subtype-specific vulnerabilities support hypothesis-driven precision therapy development in a disease where systemic treatment options remain limited.

This study represents the first comprehensive proteomic characterization of pACT and one of the largest proteomic analyses of ACT to date, with a mean of over 8,000 proteins quantified per tumor sample and more than 10,700 protein groups across the cohort. With 83 tumor samples from 72 patients, the cohort is substantial for such a rare pediatric disease. The cohort encompasses the full histological spectrum (ACA, ACX and ACC) within a richly annotated clinical framework, including endocrine phenotype, Ki67 proliferation index, vascular invasion, tumor staging, and long-term survival follow-up, with matched normal adrenal cortex specimens, enabling robust tumor-versus-normal comparisons while controlling for patient-level variation. Whole-proteome profiling enabled simultaneous interrogation of metabolic, proliferative, immune, and lineage programs, providing multidimensional biological resolution. The use of FFPE tissue, the standard archival material in clinical pathology, further supports the translational applicability of this approach.

Nevertheless, several limitations warrant consideration. The retrospective design introduces potential selection bias. Bulk proteomics averages signals across tumor and stromal compartments, limiting resolution of microenvironmental contributions^24,37^. Furthermore, mass spectrometry preferentially detects abundant proteins, potentially underrepresenting low-abundance transcription factors and signaling intermediates^22–24^. Additionally, protein abundance does not directly measure activation state^24^. Phosphoproteomic profiling would be required for precise pathway activity assessment^39^. Finally, while the five-protein classifier showed strong internal performance, it has not yet been validated in an independent cohort, which is essential before clinical implementation.

Future integration of spatial proteomics, phosphoproteomics, and multi-omic validation across independent cohorts will refine this classification framework. Importantly, prospective clinical trials incorporating molecular stratification will be required to determine whether proteome-defined subtypes can guide therapeutic decision-making and improve outcomes in pACT.

In conclusion, unsupervised proteomic profiling of 83 pACT defines four biologically and clinically distinct tumor subtypes integrating metabolic specialization, proliferative signaling, immune context, and adrenal lineage features. These proteome-defined subtypes provide superior prognostic resolution compared with histopathology alone and can be approximated by a five-protein immunohistochemistry-compatible classifier. Systematic interrogation of cluster-specific proteomes revealed distinct therapeutic vulnerabilities, including IGF-axis activation in steroidogenic subtypes and epigenetic and replication targets in high-risk tumors. This framework establishes a molecular taxonomy of pACT and a foundation for molecularly informed risk assessment and precision therapy. Prospective validation of both the classifier and the subtype-specific therapeutic vulnerabilities will be critical next steps toward clinical implementation.

## Methods

### Study design and ethical approval

We obtained the proteomic profiling cohort of pACT specimens through the German MET-Registry (n = 83 samples from n = 72 patients) including matched non-tumor adrenal controls (n = 43 specimens). Tumor specimens included ACT subtypes annotated as ACC, ACA, and adrenocortical neoplasms of uncertain malignant potential (ACX). Clinicopathological variables included sex, age at diagnosis, functional activity/endocrine phenotype, Ki67 index, Childreńs Oncology Group (COG) stage, pathological tumor (pT) stage, and follow-up parameters including death status and overall survival time (Supplementary Table S1). All samples and associated clinical data were pseudonymized.

Patient material, clinical data, and written informed consent for data and biospecimen use in scientific projects were obtained from all included patients and/or legal guardians at the time of registry enrolment, in accordance with national regulations. The MET studies were conducted in accordance with the Declaration of Helsinki and were approved by the ethics committees of the University of Lübeck (IRB 97125) and Otto-von-Guericke University Magdeburg (IRBs 174/12 and 52/22), Germany. The “Molecular basis of Pediatric Adrenocortical Tumors” (MoPACT) project (IRB 23-0351) was approved by the Ethics Committee of the Ludwig Maximilians University Munich, Germany.

### Processing of tissue samples

Tissue samples were fixed in buffered 10% formalin after surgical resection and processed into formalin-fixed, paraffin-embedded (FFPE) tissue blocks after extended fixation (typically 12–15 days). FFPE tissue was sectioned at 10 µm and mounted on adhesive glass slides (TOMO IHC Adhesive Glass Slide, Matsunami) or membrane slides (1.0 PEN, Zeiss). Representative tissue sections were deparaffinized and stained with hematoxylin and eosin for histopathological assessment. Tumor regions were inspected and marked by board certified pathologists, and the annotated tumor regions were subsequently macrodissected (scalpel-based) to enrich for tumor content prior to proteomic analysis; normal adrenal regions were selected analogously from non-tumor tissue. Macrodissection was performed from a standardized tissue area of 25 mm² per sample.

### Proteomics measurement and primary quantification

Macrodissected FFPE tissue sections were collected into a 96-well format (AFA-TUBE TPX Plate, Covaris), with edge wells left empty to minimize evaporation or temperature gradient effects. Tissue was lysed and proteins extracted from FFPE sections using a DDM-based lysis buffer, followed by Adaptive Focused Acoustics (AFA) sonication and acetonitrile-assisted antigen retrieval. Proteins were digested overnight with trypsin and LysC, and digestion was quenched by acidification to 1% TFA.

LC–MS/MS analysis was performed on a Thermo Orbitrap Astral mass spectrometer coupled to an Evosep One system (60 samples/day, 21-min gradient), equipped with a FAIMS Pro interface (−40 V). Approximately 200 ng of peptides were injected and separated on an IonOpticks Aurora Rapid C18 column at 50 °C. Raw data were analyzed using DIA-NN (v1.8.1) with a predicted spectral library from the UniProt human reference proteome, with precursor and protein group FDR controlled at 1%.

### Primary data analysis, quality control and normalization

Downstream processing was performed in R (v4.5.2). Per-sample completeness was assessed; process controls were used for QC only and excluded from analyses. Protein filtering followed a dual-filter strategy: a strict filter (≥50% detection per group) was applied for multivariate and visualization analyses; a liberal filter (≥3 non-missing values in ≥1 group) preserved group-specific proteins and was used exclusively for differential abundance testing. Intensities were log2-transformed to correct right-skew and approximate normality. Missing values in the liberally filtered matrix were imputed using a sample-wise Gaussian left-tail approach (Perseus default). For analyses requiring standardized features, row-wise Z-score normalization was applied to the strictly filtered log2 matrix.

### Bioinformatics and statistical analysis

Exploratory analyses were performed on the filtered dataset (tumor and normal; empty controls excluded). PCA (prcomp) and UMAP (umap R package; default parameters, fixed seed) were computed from row-wise Z-score-normalized protein profiles. Sample similarity was visualized as hierarchically clustered Euclidean distance heatmaps (Ward’s D2 linkage). For display heatmaps, the 500 most variable proteins [by median absolute deviation (MAD)] were selected. Differential protein abundance (tumor vs. normal) was tested using limma on the imputed, liberally filtered log2 matrix. A cell-means model (∼0 + Group) with a tumor−normal contrast was fitted, and moderated t-statistics were obtained via eBayes with robust empirical Bayes moderation. P-values were adjusted by Benjamini–Hochberg (BH) correction; significance thresholds were FDR < 0.05 and |log2 FC| ≥ 1.

Functional enrichment combined over-representation analysis (ORA) and pre-ranked gene-set enrichment analysis (GSEA). ORA was run separately on proteins with increased and decreased abundance against GO Biological Process (GO:BP), KEGG, and Reactome databases, with the full set of quantified proteins as background. For GSEA, a ranked list was generated from moderated t-statistics and tested against MSigDB Hallmark (H), C2 Canonical Pathways (C2:CP), oncogenic signatures (C6), and chemical/genetic perturbation (C2:CGP) collections. To identify the most robust biological signals, all significant results (adjusted p < 0.05) were integrated across databases using two complementary approaches: (i) a multi-source convergence analysis using ActivePathways (Brown’s method for combining correlated p-values across sources, FDR-corrected); and (ii) extraction of leading-edge proteins from all GSEA results to identify proteins recurrently driving enrichment across multiple independent gene sets. A composite priority score was computed to rank biological themes by their breadth of cross-database support, effect size, and mechanistic centrality.

Supervised tumor subgroup analyses were performed on samples classified as ACC, ACA, or ACX. An omnibus limma moderated F-test identified discriminating proteins, followed by pairwise contrasts. A limma trend model with ordinal coding (ACA = 1, ACX = 2, ACC = 3) tested for monotonic abundance changes along the putative malignancy spectrum. Directed GSEA (ACC vs. ACA) was performed against GO:BP and Reactome.

Unsupervised tumor stratification used consensus clustering (ConsensusClusterPlus; Ward’s D2, Euclidean distance, k = 2–6, 1,000 iterations, 80% sample resampling, all features) on the full tumor Z-score matrix. Non-negative matrix factorization (NMF) rank surveys (ranks 2–6, 30 runs per rank, Brunet algorithm) independently validated the optimal k. Cluster-defining proteins were identified by limma one-versus-rest contrasts (BH-adjusted P < 0.05, log2 FC > 0.5). Cluster associations with clinical variables were tested by Fisher’s exact (categorical) or Kruskal–Wallis (continuous) tests. Overall survival was analyzed by Kaplan–Meier estimation with log-rank testing.

A minimal immunohistochemistry (IHC) panel was derived by greedy forward selection with leave-one-out cross-validated nearest-centroid classification (panel size 5–20; significance assessed by 1,000-permutation test).

Adrenal zonation was inferred using curated marker panels for zona glomerulosa (ZG), zona fasciculata (ZF), and zona reticularis (ZR) (Supplementary Table S2); per-sample zone scores (mean Z-score of detected markers) were converted to proportions via softmax transformation and visualized as ternary diagrams. The analyses which were aimed at clinical translation comprised: (i) drug repurposing via CMap-proxy GSEA (C2:CGP perturbation signatures); (ii) druggable proteome annotation against a curated database of ∼360 targets across 20 pharmacological families (Supplementary Table S3); (iii) kinase activity inference by single sample (ss)GSEA on KEGG/Reactome signaling pathways; (iv) pathway-specific analyses of IGF2/IGF1R, Wnt/β-catenin, and TP53 signaling (Supplementary Tables S4–S6); (v) immune microenvironment estimation using curated cell-type markers (Supplementary Table S7); (vi) fetal-versus-adult adrenal signature scoring (Supplementary Table S8); and (vii) metabolic vulnerability profiling (ssGSEA on KEGG/Hallmark metabolic pathways).

Detailed descriptions of all analyses including the key packages used in R v4.5.2 (R Core Team, 2025) are available in the Supplementary Methods and Supplementary Table S9. All tests were two-sided; BH correction at FDR 5% was applied unless stated otherwise.

## Data Availability

All data produced in the present study are available upon reasonable request to the authors

## Acknowledgment

We gratefully acknowledge the patients and their families for their trust and participation in this study. We thank the physicians, study nurses, and clinical staff at the contributing institutions for their commitment to patient care and for providing clinical information and biospecimens to the German MET Registry.

We are indebted to the technical staff of the participating pathology and proteomics laboratories for expert sample processing, mass spectrometry measurements, and data management. We particularly thank Lena Reichl and Maria Kling for excellent technical assistance.

## Declaration of generative AI and AI-assisted technologies in the manuscript preparation process

During the preparation of this work the authors used OpenEvidence, Nature Research Assistant, ChatGPT, Claude AI and perplexity.ai in order to brainstorm and enhance readability of sections of the text. After using these tools, the authors reviewed and edited the content as needed and take full responsibility for the content of the published article.

## Authorś contributions

R. Claus: Conceptualization, Data Curation, Formal Analysis, Funding Acquisition, Investigation, Methodology, Project Administration, Resources, Software, Supervision, Visualization, Writing – Original Draft Preparation; A. Metousis: Data Curation, Formal Analysis, Investigation, Methodology, Validation, Writing – Review & Editing; V.E. Fincke: Validation, Writing – Review & Editing; M. Kunstreich: Data Curation, Investigation, Resources, Writing – Review & Editing; S.A. Wudy: Supervision, Validation, Writing – Review & Editing; E. Jüttner: Investigation, Resources, Writing – Review & Editing; J. Pons-Kühnemann: Formal Analysis, Writing – Review & Editing; C. Vokuhl: Investigation, Resources, Writing – Review & Editing; M.C. Frühwald: Writing – Review & Editing; C. Röcken: Investigation, Resources, Writing – Review & Editing; L. Schweizer: Investigation, Supervision, Writing – Review & Editing; P.D. Johann: Supervision, Validation, Writing – Review & Editing; A. Redlich: Data Curation, Funding Acquisition, Investigation, Project Administration, Resources, Writing – Review & Editing; M. Mann: Investigation, Methodology, Resources, Writing – Review & Editing; M. Kuhlen: Conceptualization, Data Curation, Funding Acquisition, Investigation, Methodology, Project Administration, Resources, Supervision, Validation, Writing – Original Draft Preparation

## Supplementary Figure legends

**Supplementary Figure S1.**
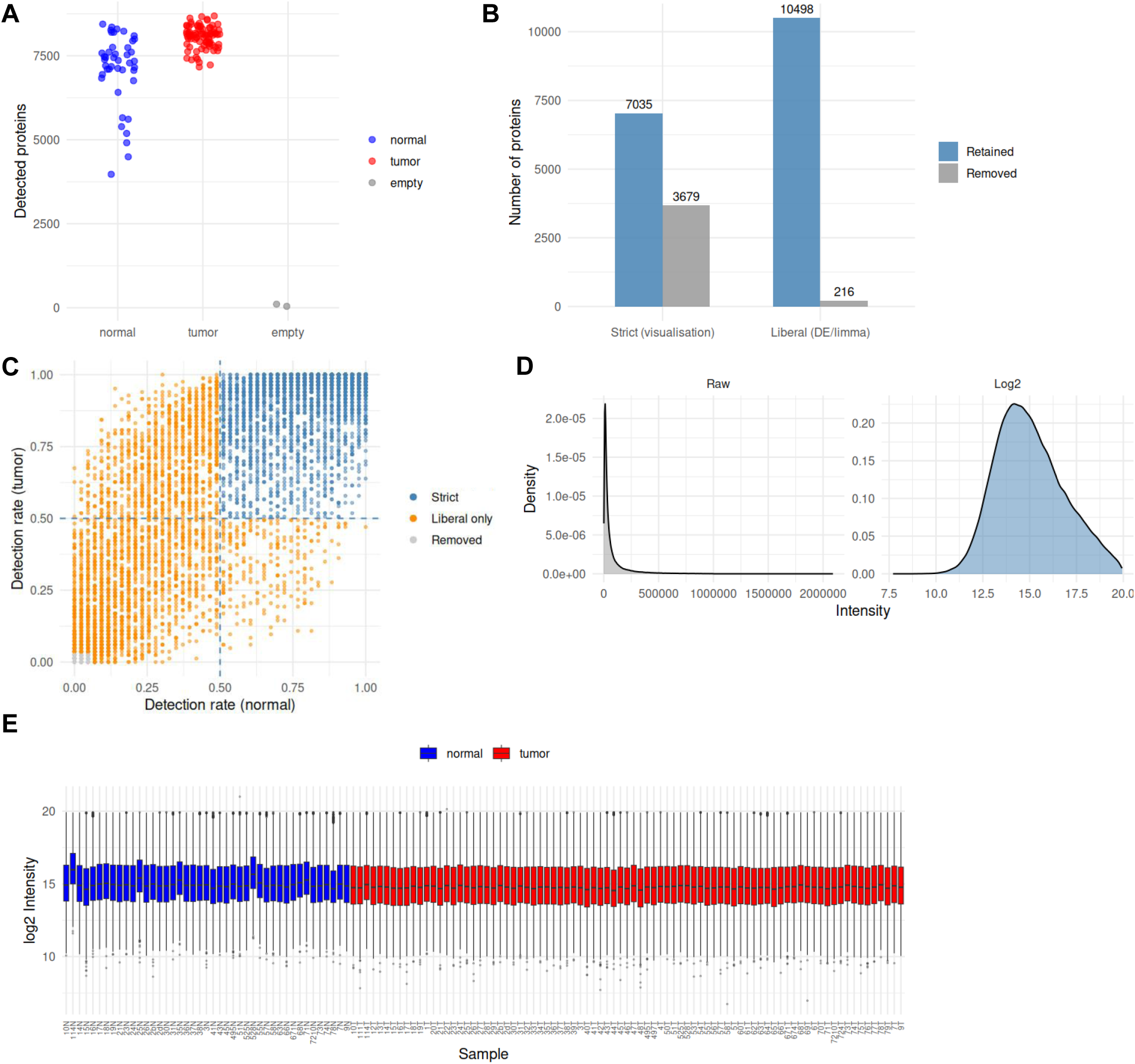
Protein filtering, quality control, and data transformation. (A) Per-sample detected protein group counts across all three sample types (normal, tumor and empty controls), derived from the unfiltered raw expression matrix. Empty control runs yield near-zero identifications, confirming measurement specificity. (B) Protein group retention and removal under the two parallel filtering strategies. The strict filter (≥50% detection in each group) retained 7,035 proteins for visualization; the liberal filter (≥3 valid values in at least one group) retained 10,498 proteins for differential abundance analysis. Counts of retained and removed proteins are shown for each strategy. (C) Per-protein detection rate in tumor versus normal samples, colored by filter classification: strict (blue; passes both filters), liberal only (orange; retained for differential abundance only; group-specific proteins), and removed (grey; excluded from all analyses). Dashed lines indicate the 50% detection threshold applied by the strict filter. (D) Distribution of protein intensities before and after log2 transformation, shown as density plots. The raw distribution is strongly right-skewed; log2 transformation yields a near-symmetric, bell-shaped distribution centered around 15. (E) Per-sample log2 intensity boxplots for all normal (blue) and tumor (red) samples, derived from the strictly filtered, log2-transformed matrix (7,035 proteins; observed values only, no imputation). Distributions are consistent across samples, confirming the absence of systematic per-sample intensity biases.

**Supplementary Figure S2.**
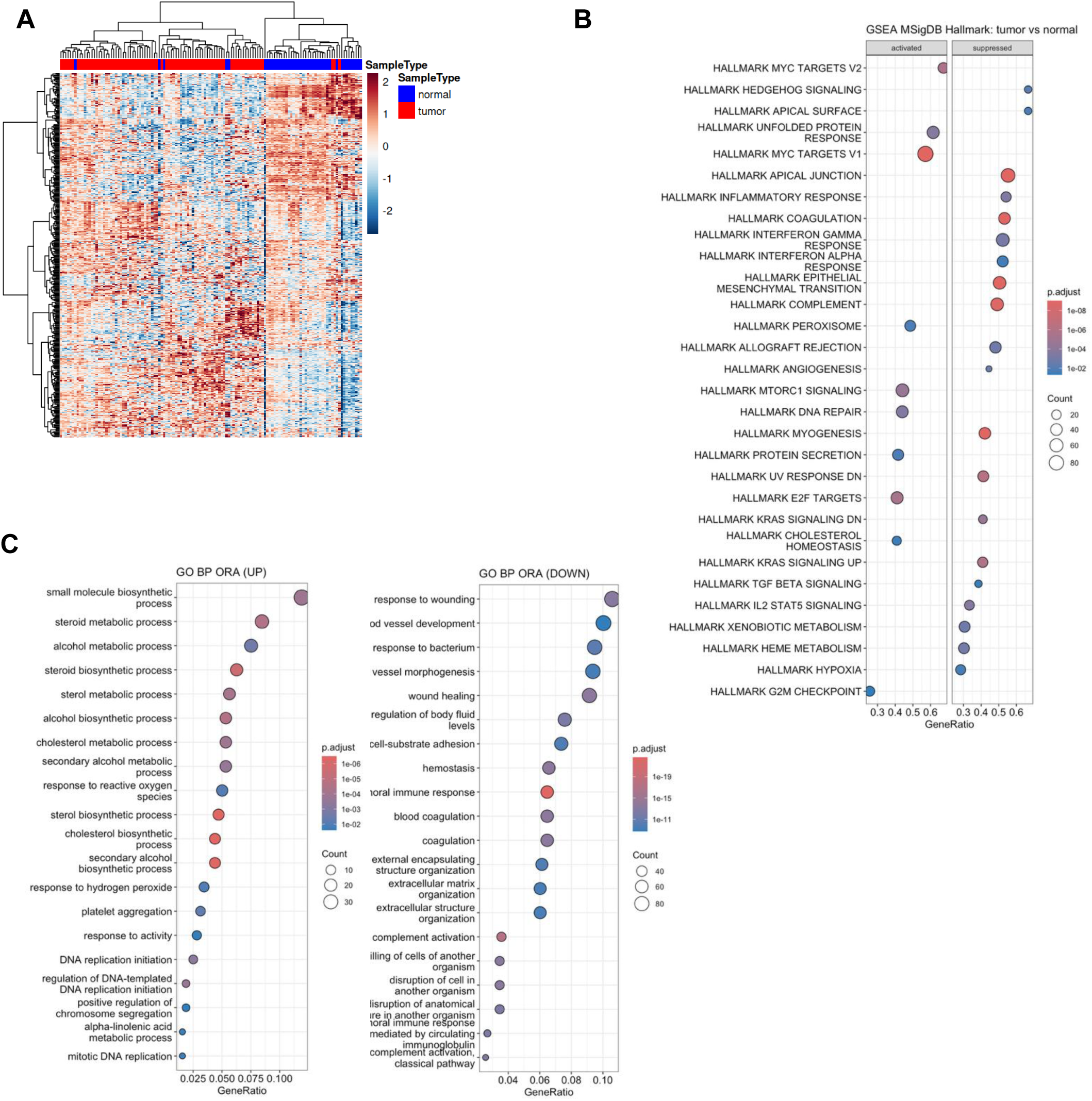
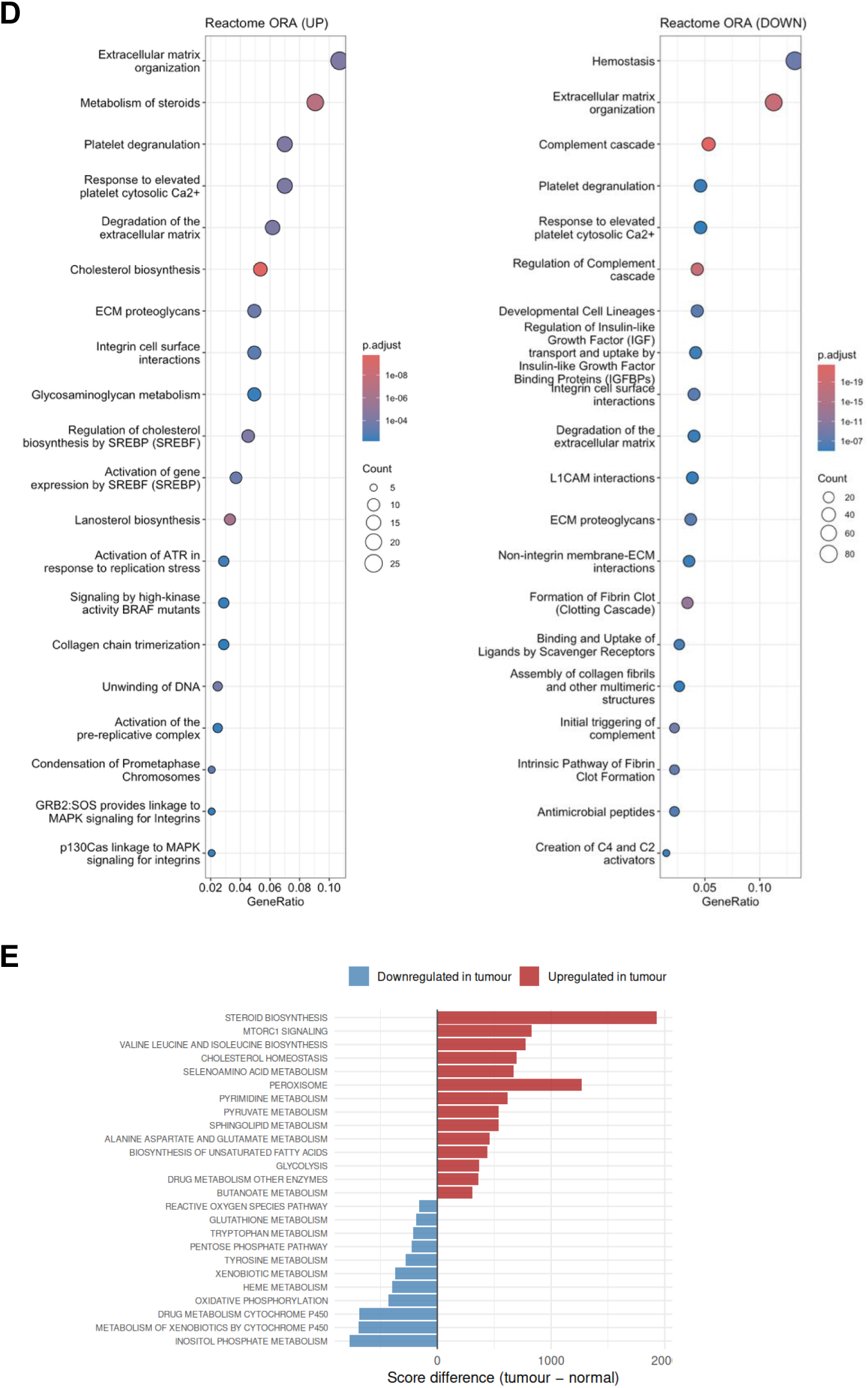
Additional unsupervised clustering and pathway enrichment analyses. Panel A uses the strictly filtered matrix (7,035 proteins); panels B–E use the liberally filtered, imputed matrix (10,498 proteins). (A) Heatmap of the 500 most variable proteins. Proteins were selected by variance and visualized after per-protein Z-score normalization; hierarchical clustering of both proteins and samples used Ward’s method. Annotation bar indicates sample group (tumor vs. normal). (B) MSigDB Hallmark gene set enrichment analysis (GSEA) for the tumor vs. normal comparison, performed on a ranked list of moderated t-statistics. Significantly enriched Hallmark gene sets are shown, with facets indicating enrichment direction (activated vs. suppressed in tumors). Dot size reflects contributing protein count; color indicates adjusted p-value. (C) GO BP pathway ORA for metabolic pathways, stratified by direction of change (FDR < 0.05, |log2 FC| ≥ 1). GeneRatio on the x-axis; dot size proportional to Count; color indicates adjusted p-value. (D) Reactome pathway ORA for signaling and cellular interaction pathways, stratified by direction of change. Display parameters as in panel C. (E) Per-pathway single-sample GSEA (ssGSEA) score differences (tumor minus normal) for KEGG metabolic pathways. Bars are colored by enrichment direction (red: increased in tumors; blue: decreased). Pathways are ranked by absolute score difference.

**Supplementary Figure S3.**
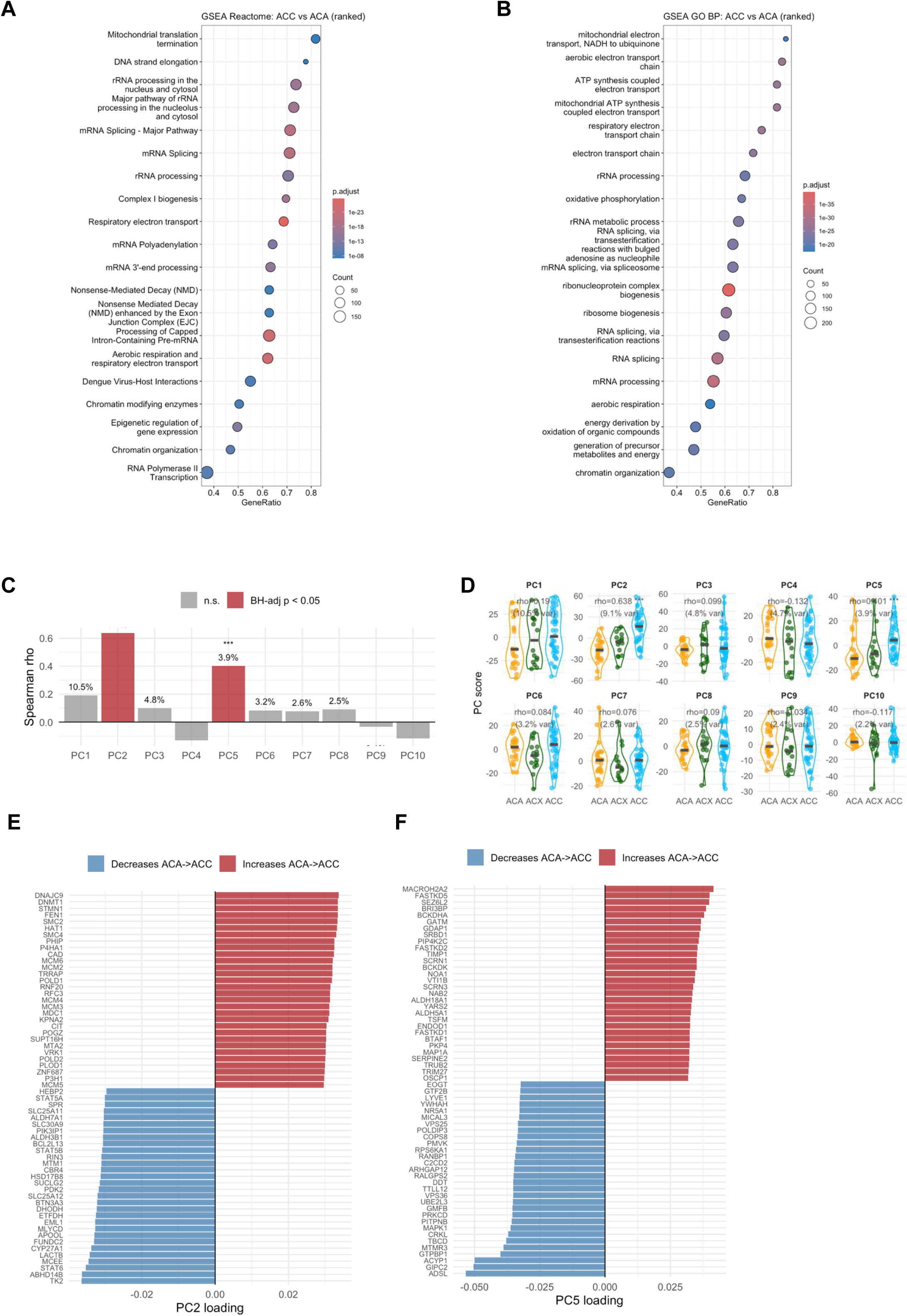
Gene set enrichment analysis for the ACC vs. ACA (tumor-only analysis). All panels use the strictly filtered, log2-transformed tumor-only matrix (7,035 proteins). (A, B) GSEA dot plot for Reactome and GO BP pathways. Dot size reflects the number of contributing proteins (Count); color indicates adjusted p-value. Facets or color coding distinguish gene sets activated in ACC versus those suppressed in ACC relative to ACA. (C) Spearman rank correlation between PC scores (PCs 1–10) and ordinal subgroup coding (ACA = 1, ACX = 2, ACC = 3). Bar height indicates rho; color denotes significance (red: BH-adjusted p < 0.05; grey: not significant). Percentages show variance explained per PC. (D) Distribution of PC scores across diagnostic subgroups for PCs 1–10. Each point represents one tumor; horizontal crossbars indicate group medians. Rho and variance explained are annotated per panel. (E) Top 30 protein loadings on PC2 in each direction. Red bars: proteins whose abundance increases along the ACA-to-ACC axis; blue bars: proteins that decrease. (F) Top 30 protein loadings on PC5, displayed as in (E).

**Supplementary Figure S4.**
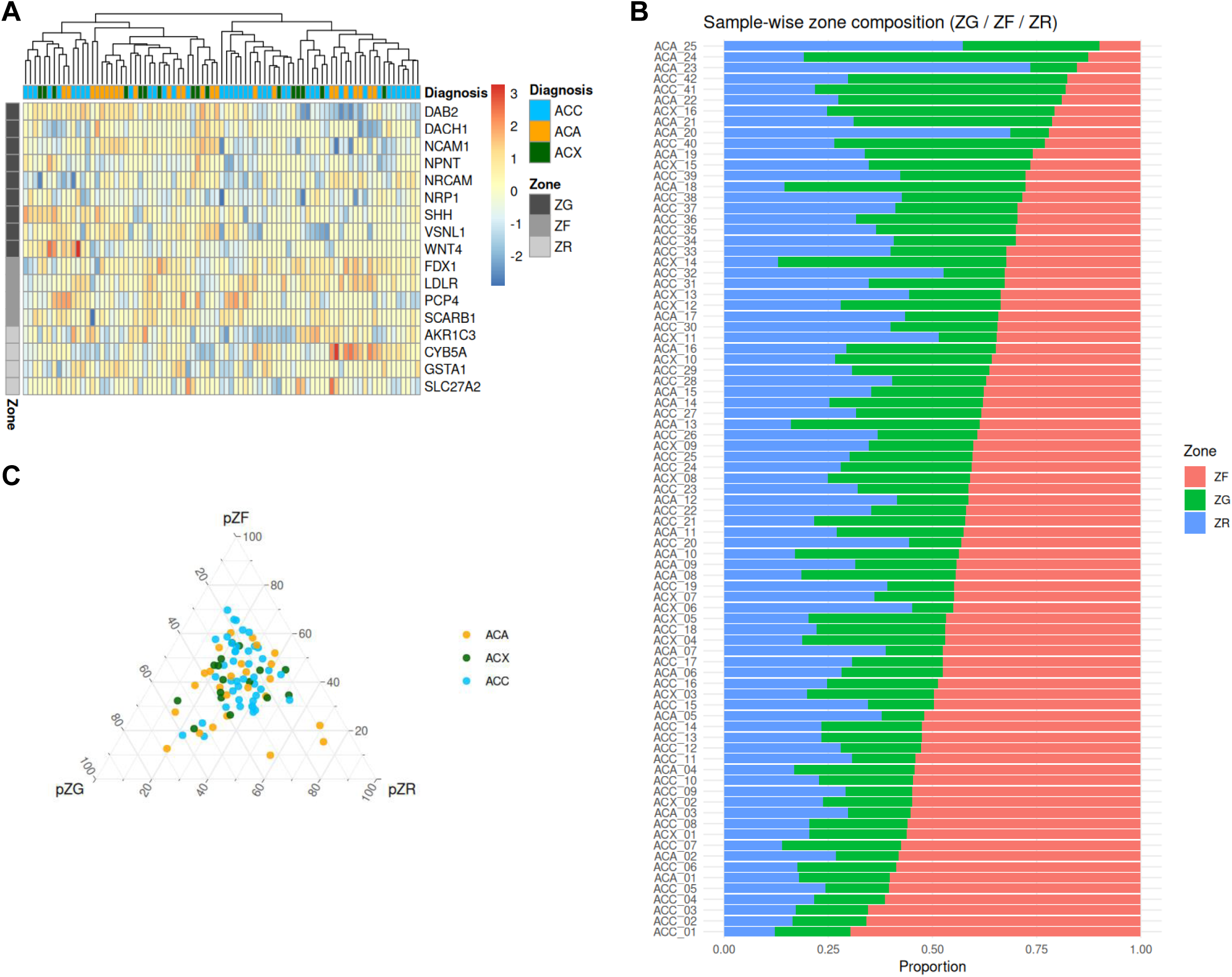
Adrenal zonation marker analysis (tumor-only). All panels use the strictly filtered, Z-score-normalized tumor-only matrix. Zone scores were computed as the mean Z-score of the best-detected marker per gene symbol (minimum 2 markers required per zone). (A) Heatmap of detected ZG, ZF, and ZR zone markers. Rows are ordered by zone block (ZG/ZF/ZR) without row clustering; samples are hierarchically clustered by Ward’s method. Annotation bars indicate diagnosis (ACC/ACA/ACX) and zone block assignment. (B) Sample-wise stacked bar chart showing proportional ZG/ZF/ZR zone composition. Zone scores were converted to proportions via softmax transformation; bars sum to 1 per sample. Samples are ordered by diagnosis (ACA, ACX, ACC). (C) Ternary plot of sample-wise zone compositions (pZG, pZF, pZR), with each vertex representing 100% contribution of one zone. Points are colored by diagnosis (ACC: blue, ACA: orange, ACX: green).

**Supplementary Figure S5.**
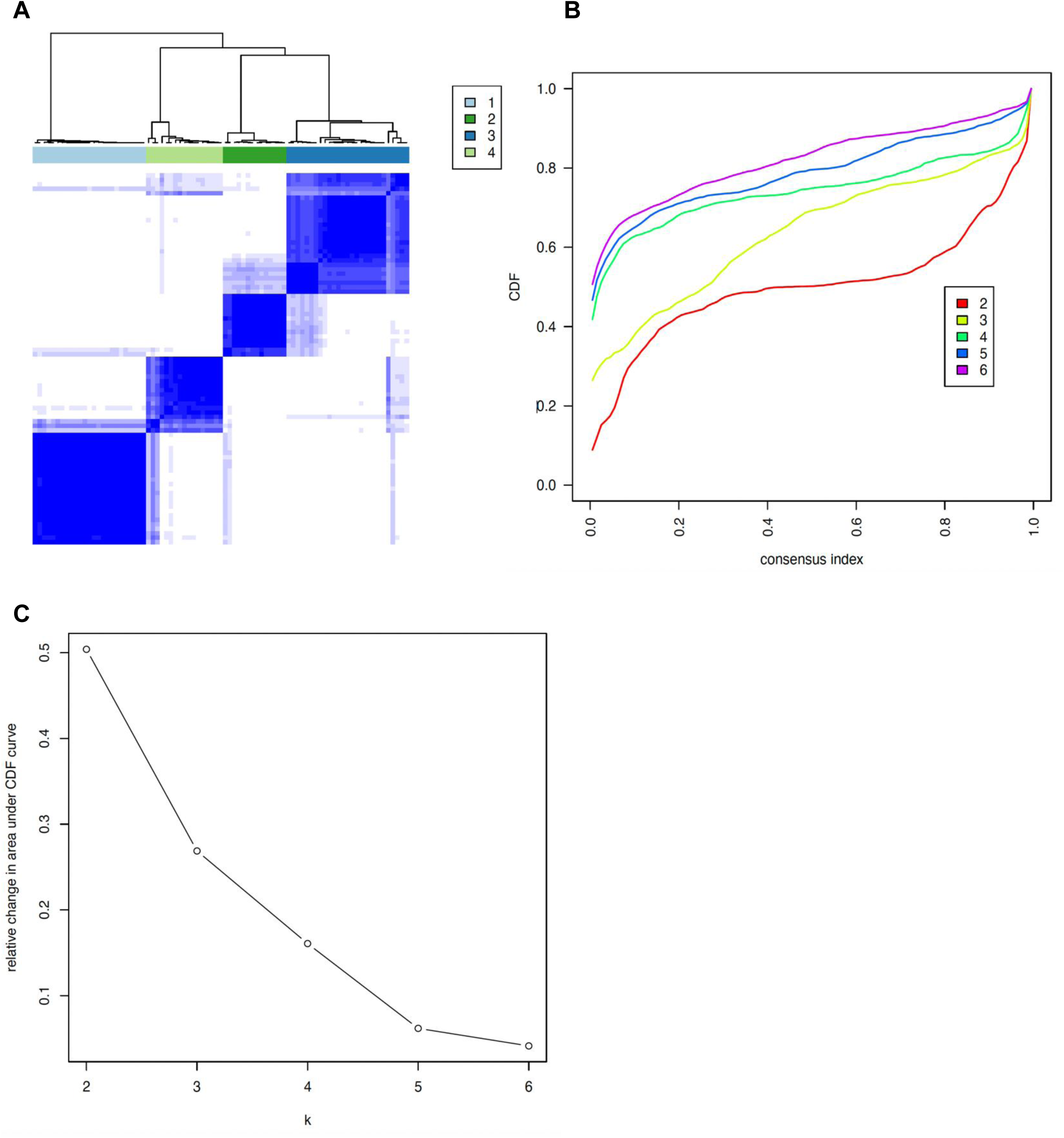
Consensus clustering quality metrics supporting the k = 4 solution. (A) Consensus clustering heatmap for k = 4, showing the consensus matrix across all tumor samples (1,000 resamplings). Color intensity represents the proportion of resampling runs in which two samples were assigned to the same cluster; dark diagonal blocks indicate high within-cluster stability. (B) Cumulative distribution function (CDF) of consensus scores for k = 2–6. A flat CDF indicates stable cluster assignments; k = 4 shows a marked improvement over lower k values. (C) Relative change in area under the CDF curve (delta area) across k values. The largest incremental gain in cluster stability occurs at k = 4, after which additional clusters produce diminishing returns, supporting selection of k = 4 as the optimal partition.

**Supplementary Figure S6.**
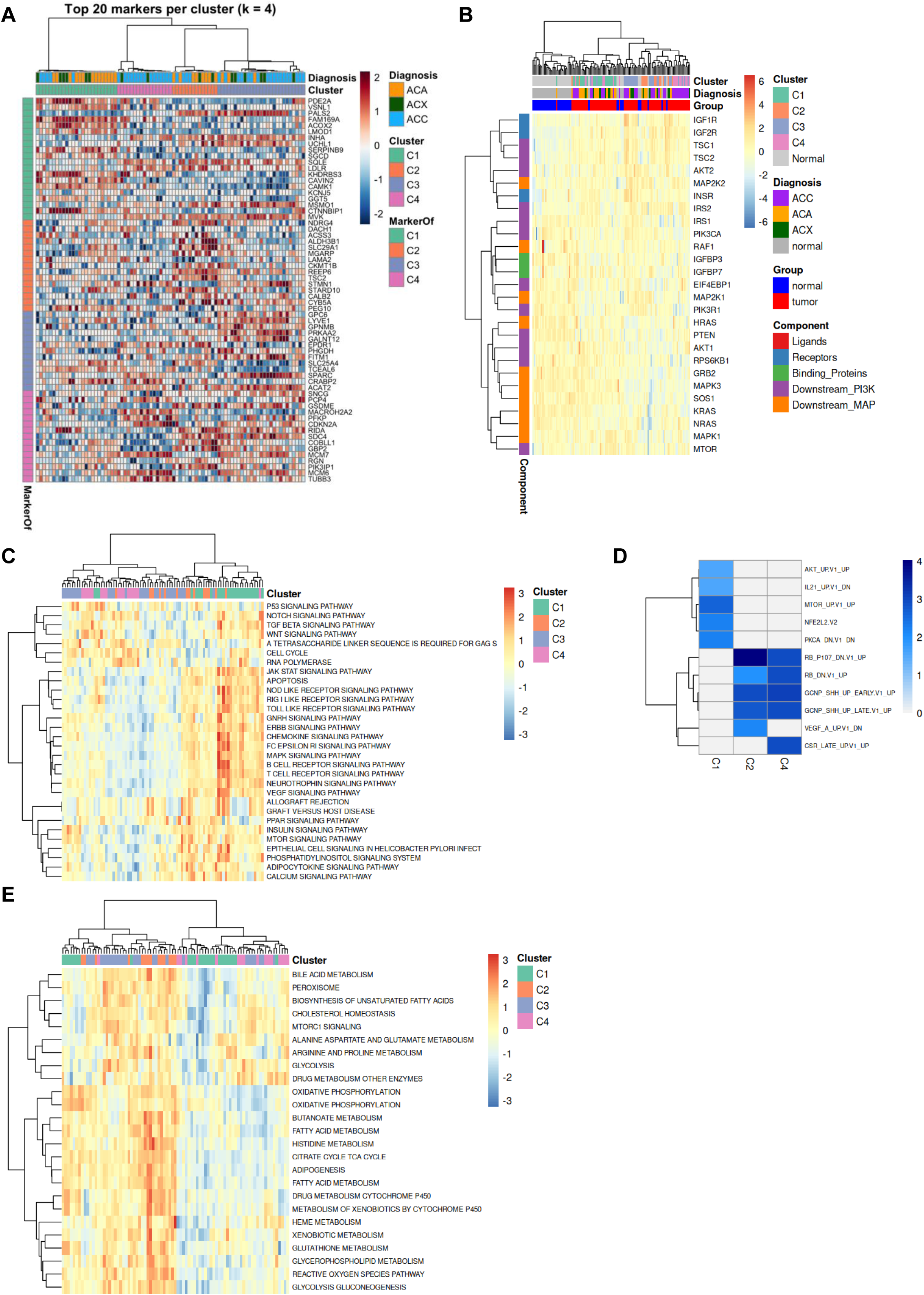
Cluster-specific marker protein abundance and pathway enrichment detail. All panels derive from the strictly filtered, log2-transformed tumor-only matrix; panel A uses a one-vs-rest limma contrast per cluster (empirical Bayes, BH-adjusted). Panel B includes matched normal samples for comparison. (A) Heatmap of top cluster-defining marker proteins (top 20 per cluster by absolute log2 FC, FDR < 0.05, |log2 FC| ≥ 0.5). Z-score-normalized values are shown; annotation bars indicate proteome cluster, diagnosis and marker assignment (“MarkerOf”). (B) Heatmap of IGF signaling pathway component proteins across clustersand matched normal samples. Annotation bars indicate cluster, diagnosis, and group (tumor/normal); row annotation indicates signaling component class. (C) KEGG signaling pathway over-representation analysis (ORA) heatmap per cluster (cluster vs. rest; FDR < 0.05). Dot size proportional to gene ratio; color indicates adjusted p-value. (D) Gene set enrichment heatmap for oncogenic signature gene sets (MSigDB C6) per cluster. (E) KEGG metabolic pathway enrichment score heatmap per cluster, highlighting cluster-specific metabolic program differences.

**Supplementary Figure S7.**
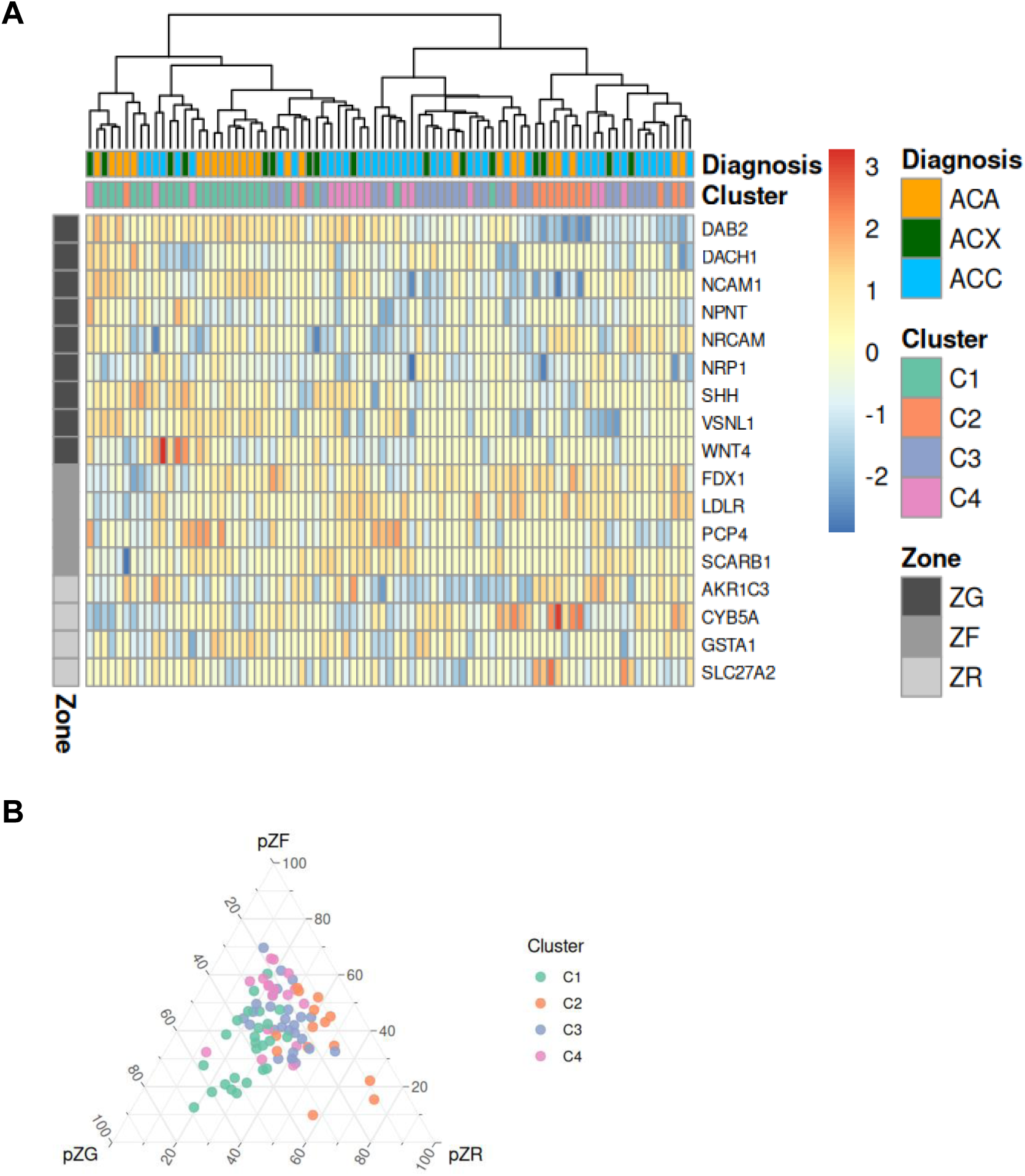
Ternary plot for adrenal zonation and marker heatmap across proteome clusters. (A) Ternary plot of sample-wise adrenal zone compositions (pZG, pZF, pZR) stratified by proteome cluster. Each vertex represents 100% contribution of one zone; scores were converted to proportions via softmax transformation. (B) The panel uses the strictly filtered, Z-score-normalized tumor-only matrix. Rows are ordered by zone block (ZG/ZF/ZR) without row clustering; samples are hierarchically clustered by Ward’s method. Annotation bars indicate diagnosis (ACC blue, ACA orange, ACX green), proteome cluster (C1–C4), and zone block assignment (ZG/ZF/ZR).

**Supplementary Figure S8.**
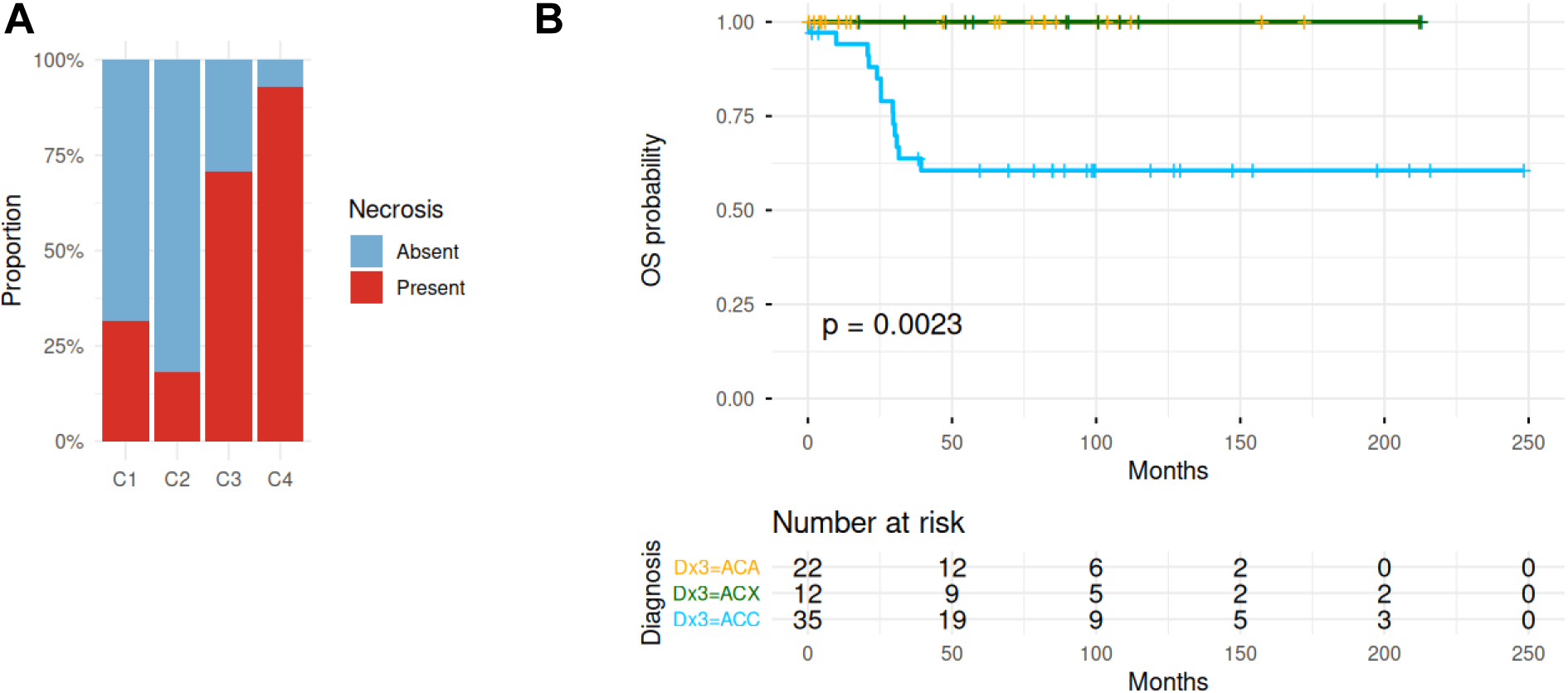
Additional clinical associations. (A) Necrosis status across proteome clusters, shown as stacked proportion bars (Fisher’s exact p < 0.0001). Necrosis was present in 32% of C1, 18% of C2, 71% of C3, and 93% of C4 tumors. (B) Kaplan–Meier overall survival curves stratified by histological diagnosis (ACA, ACX, ACC) for comparison with the cluster-based survival analysis in Figure 5I (log-rank p = 0.0023). Numbers at risk are shown below the plot.

**Supplementary Figure S9.**
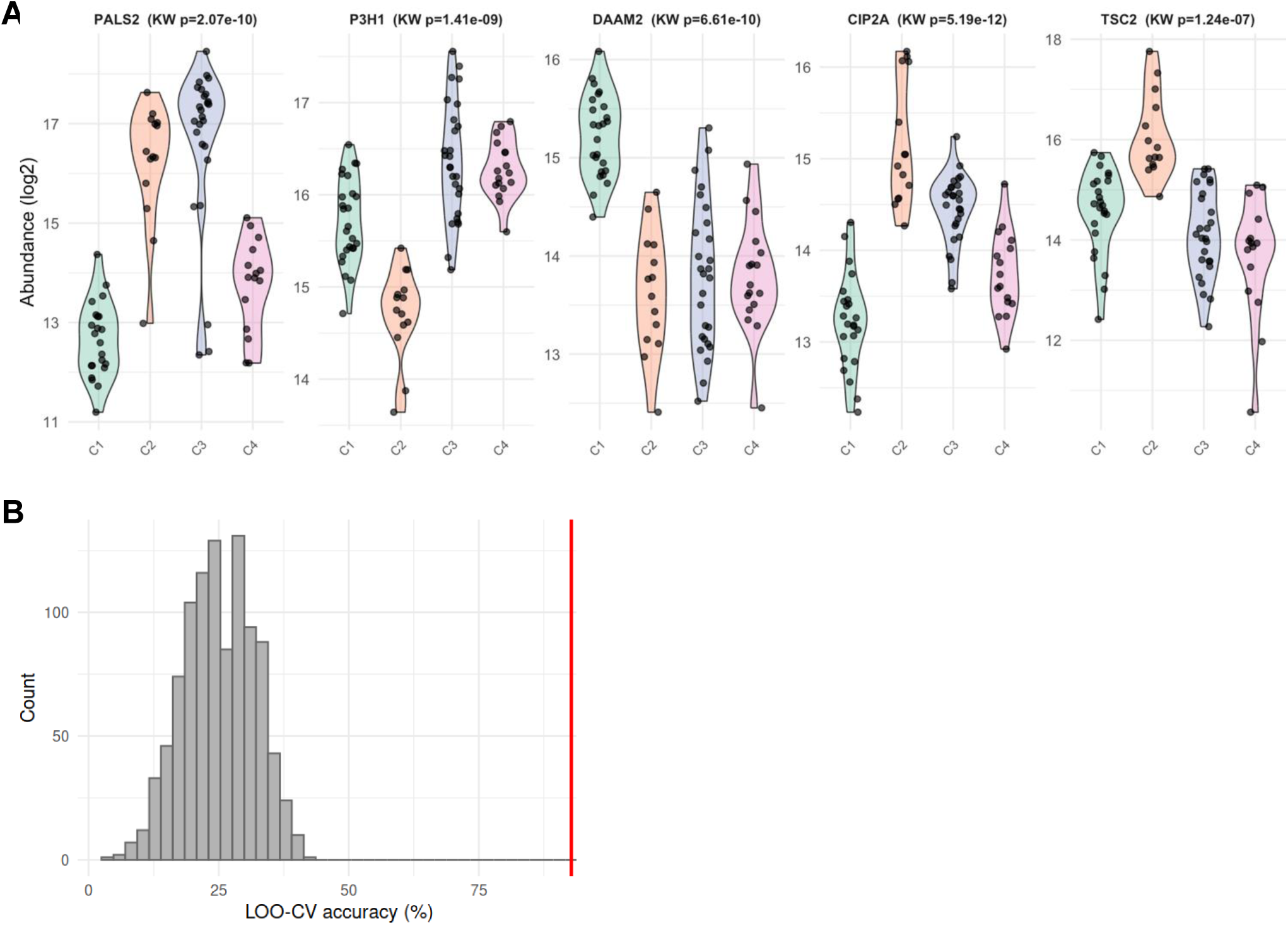
Individual classifier protein abundance and permutation test. (A) Violin plots of log2 abundance per cluster for each classifier panel protein: PALS2, P3H1, (DAAM2, CIP2A, TSC2. Kruskal-Wallis p < 0.001 for all. (B) Distribution of LOO-CV accuracies from 1,000 cluster-label permutations (grey histogram); red line indicates observed accuracy (92.8%; permutation p < 0.001).

**Supplementary Figure S10.**
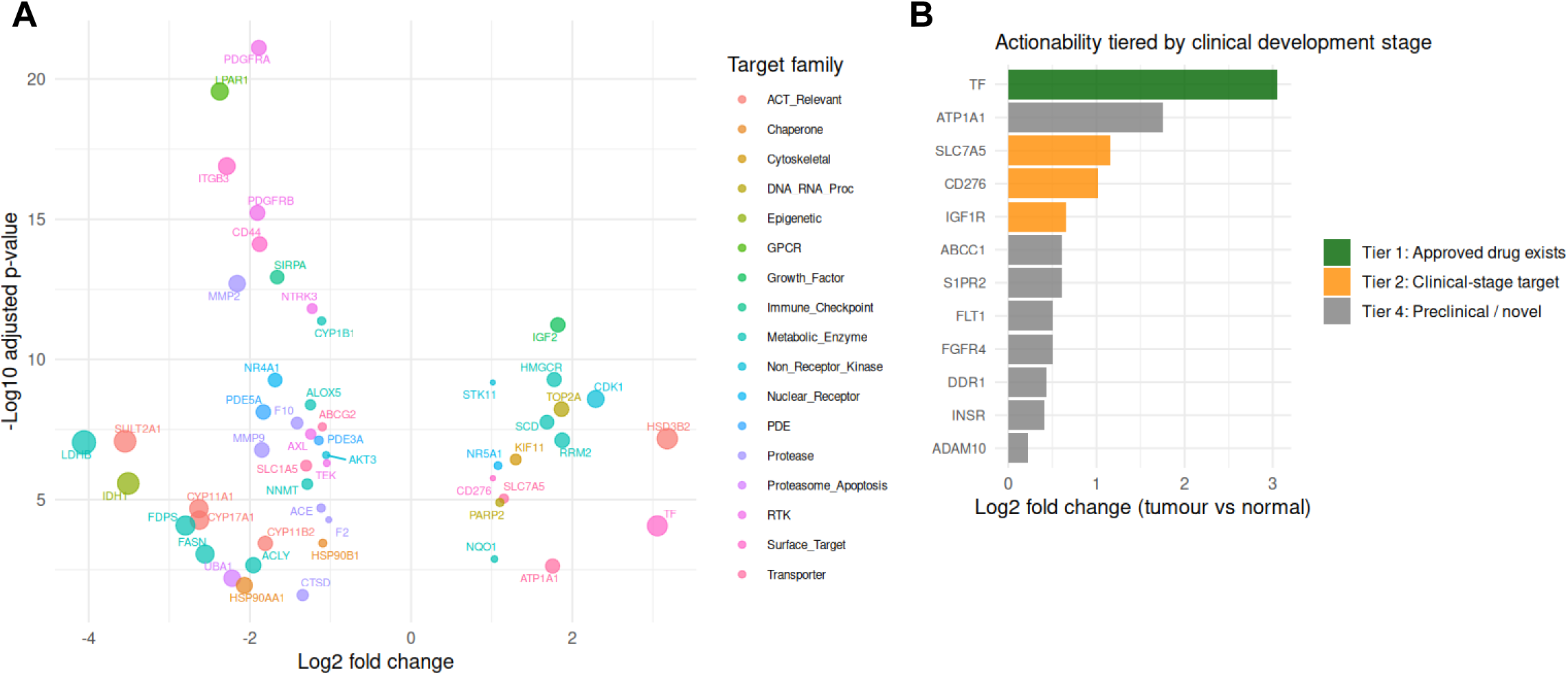
Extended therapeutic target actionability analysis. (A) Volcano plot of tumor vs. normal differential abundance for proteins within the curated druggable target set, colored by target family (RTK, kinase, GPCR, protease, etc.). Dot size reflects statistical significance (−log10 adjusted p-value). Proteins exceeding significance and fold-change thresholds are labeled. (B) Bar chart of actionable targets stratified by clinical development stage (Tier 1–4) and cluster, showing log2 fold change (tumor vs. normal) for top candidates per target class. Color indicates actionability tier.

