## Supplementary material for "A proteome-based classification of pediatric adrenocortical tumors links functional tumor states to clinical outcome and therapeutic vulnerabilities": Suppl Material

### Supplementary Methods

#### Proteomics sample preparation

Macrodissected FFPE tissue sections were collected into a 96-well AFA-TUBE TPX Plate (Covaris), with edge wells left empty to minimize evaporation and temperature gradient effects. Tissue was lysed in 42 µL lysis buffer (70 mM TEAB in MS-grade water with 0.013% DDM) by incubation at 90 °C for 60 min, followed by cooling to room temperature. Samples were sonicated using Adaptive Focused Acoustics (AFA) in 96-well format (LE220-plus, Covaris; 300 s per column; peak power 450, duty factor 50%, 200 cycles, average power 225). Following sonication, 7 µL of 80% acetonitrile in MS-grade water was added and samples were incubated at 75 °C for 60 min. Proteins were digested overnight at 37 °C with 1 µg trypsin and 1 µg LysC at 1,200 rpm shaking. Digestion was quenched the following day by acidification to a final concentration of 1% TFA.

#### LC–MS/MS acquisition parameters

Approximately 200 ng of digested and acidified peptides were loaded onto C18 EvoTips Pure according to the manufacturer's instructions. The FAIMS Pro interface was operated at a compensation voltage of −40 V. Peptides were separated on an IonOpticks Aurora Rapid 8×150 C18 column (80 mm × 0.15 mm) maintained at 50 °C. Full MS scans were acquired over 380–980 m/z at an Orbitrap resolution of 240,000 with a normalized AGC target of 500%. For MS/MS scans, ions from 4-Th-wide isolation windows spanning the 380–980 m/z range were accumulated for a maximum injection time of 5 ms and fragmented by HCD at a normalized collision energy of 25%.

#### DIA-NN search parameters

Raw data were analyzed using DIA-NN (v1.8.1) with a predicted spectral library generated from the UniProt human reference proteome (UP000005640_9606 and UP000005640_9606_additional). Precursor charge states ranged from 1 to 4; precursor mass range was set to 300–1,300 m/z; fragment ion mass range to 100–1,800 m/z; and peptide length from 7 to 35 amino acids. Mass accuracy and MS1 mass accuracy were fixed at 17 and 13 ppm, respectively, and the scan window was set to 7. Heuristic protein grouping was enabled (--relaxed-prot-inf). Library generation used the "Smart profiling" setting with retention time profiling enabled. An empirical spectral library was generated from the DIA data and used to reanalyze all runs in a two-pass workflow.

**Quality control, transformation, and imputation**

Per-sample completeness was calculated as the fraction of protein groups with non-missing intensity values; process controls ("empty" injections) were included in QC summaries but excluded from all subsequent analyses. Per-sample skewness was computed at each transformation stage (raw, log2, Z-score) to confirm that log2 transformation adequately symmetrized intensity distributions prior to statistical modelling. For the liberally filtered matrix, missing values were imputed using a sample-wise Gaussian left-tail approach identical to the Perseus default: for each sample, imputed values were drawn from $N(\mu - 1.8\sigma, (0.3\sigma)^2)$, where $\mu$ and $\sigma$ are the mean and standard deviation of the observed log2 intensities in that sample, placing imputed values in the left tail to represent proteins near or below the detection limit. Imputation quality was verified by density-plot comparison of observed versus imputed value distributions.

**Exploratory dimensionality reduction**

PCA was computed on the transposed Z-score matrix (samples as rows) without additional scaling (*scale.* = FALSE, as the data were already Z-score-normalized per protein). UMAP was run with default parameters (*n_neighbors* = 15, *min_dist* = 0.1) using the *umap* R package (McInnes *et al.*, 2018), with a fixed random seed (42). Sample-to-sample Euclidean distances were computed on Z-scored profiles and visualized as hierarchically clustered heatmaps (Ward's D2 linkage, *heatmap*). Heatmaps of the top 500 most variable proteins (ranked by MAD) were annotated with group assignment and clinical covariates.

**Differential abundance analysis**

The imputed, liberally filtered $\log_2$ abundance matrix served as input for differential testing. A cell-means design matrix (~0 + Group) was fitted per protein via *lmFit()*, and a tumor−normal contrast was evaluated with *contrasts.fit()*. Moderated t-statistics, $\log_2$ fold changes, and p-values were computed via *eBayes()* with *robust = TRUE*, which employs a heavier-tailed prior distribution to down-weight the influence of outlier proteins on the global variance estimate (Phipson *et al.*, 2016). Multiple testing was controlled by BH correction. Proteins meeting FDR < 0.05 and $|\log_2 FC| \geq 1$ were classified as significantly differentially abundant. Volcano plots labelled the 15 most significant increased and 15 most significant decreased proteins by adjusted *P*-value.

**Functional enrichment analyses**

**Over-representation analysis (ORA).** Increased and decreased protein lists were analyzed separately against Gene Ontology Biological Process (GO:BP; *enrichGO*, SYMBOL key type),

KEGG (*enrichKEGG*, ENTREZ key type), and Reactome (*enrichPathway* via ReactomePA, ENTREZ key type). Gene symbol-to-ENTREZ ID mapping was performed using *bitr()* from *clusterProfiler* with the *org.Hs.eg.db* annotation database. The full set of quantified proteins with valid gene symbols served as the background universe. BH adjustment was applied at $q < 0.05$ for both p-value and q-value cutoffs. Disease-association enrichment was additionally performed using Disease Ontology (*enrichDO*) and DisGeNET (*enrichDGN*) via the DOSE package, using ENTREZ IDs.

**Gene-set enrichment analysis (GSEA).** A pre-ranked gene list was generated from the moderated t-statistic of the tumor-versus-normal limma contrast, providing a variance-stabilized ranking metric. When multiple protein groups mapped to one gene symbol, the entry with the largest absolute t-statistic was retained to avoid redundancy while preserving the most informative measurement. The sorted gene list was tested via *clusterProfiler::GSEA()* against three MSigDB collections obtained via *msigdbr*: Hallmark (H; 50 gene sets summarizing well-defined biological processes), oncogenic signatures (C6; gene sets from oncogene/tumor-suppressor perturbation experiments), and chemical/genetic perturbation signatures (C2:CGP; ~3,400 gene sets from published studies). Gene-set size was bounded at 10–500 members, with an epsilon of $10^{-10}$ for p-value computation. BH correction was applied to all gene-set p-values.

**Leading-edge gene analysis and pathway clustering.** For each GSEA result reaching statistical significance (Benjamini–Hochberg adjusted $p < 0.05$), the leading-edge subset was extracted from the core_enrichment field of the *clusterProfiler* output. Leading-edge genes represent the subset of a gene set that contributes most to the enrichment score and lie at the point where the running enrichment statistic reaches its maximum deviation; they were interpreted as the proteins primarily driving the enrichment signal of each pathway. Leading-edge genes from all significant GSEA results (Hallmark, Reactome, C2:CP, C6, C2:CGP) were combined into a unified pathway–gene table. For each gene, the number of distinct significant pathways in whose leading edge it appeared (n_pathways) was counted, providing a recurrence measure across independent gene sets. Genes with high recurrence were considered functional hubs. To reduce redundancy among pathways, pairwise Jaccard similarity between leading-edge gene sets was computed, and pathways with Jaccard index above a moderate overlap threshold were connected in a graph. Community detection (Louvain algorithm) was then applied to cluster highly overlapping pathways into non-redundant functional themes, and one representative pathway per cluster (the pathway with the largest absolute normalized enrichment score, NES) was retained for visualization and reporting.

**Multi-source convergence and priority scoring of biological themes.** To prioritize biologically coherent themes supported across multiple enrichment methods and databases, all significant pathway results (adjusted $p < 0.05$) from GSEA (Hallmark, Reactome, C2:CP, C6, C2:CGP) and ORA (GO:BP, KEGG, Reactome, Disease Ontology, DisGeNET) were pooled into a single table annotated with pathway identifier, source database/method, adjusted p-value, and NES where applicable. Each pathway was assigned to one or more broad biological themes (e.g., "PI3K/AKT/mTOR signalling", "Cell cycle", "Fatty acid metabolism", "Interferon signalling") by keyword-based matching against pathway names; themes were defined a priori and a pathway could map to multiple themes.

For each theme, three quantitative metrics were derived: (i) the number of independent analysis sources with at least one significant pathway mapping to that theme (N_sources), reflecting cross-database/method convergence; (ii) the mean absolute NES across all significant GSEA pathways assigned to the theme, reflecting effect size; and (iii) the median recurrence of leading-edge genes associated with the theme (median number of pathways in which a theme's leading-edge genes appeared), reflecting mechanistic centrality of the driving proteins. These three components were converted to z-scores across themes and combined into a heuristic composite priority score:

$$\text{Priority score} = 0.4 \cdot z(N_{\text{sources}}) + 0.3 \cdot z(|\,\bar{\text{NES}}\,|) + 0.3 \cdot z(\tilde{f}_{\text{gene}}),$$

where $z(\cdot)$ denotes z-transformation across all themes, $|\,\bar{\text{NES}}\,|$ is the mean absolute NES per theme, and $\tilde{f}_{\text{gene}}$ is the median leading-edge gene recurrence for that theme. This score is dimensionless and centered around zero; higher values indicate themes that are supported by multiple independent databases/methods, show large absolute enrichment effects, and are driven by recurrent leading-edge hub proteins. Because the weights are heuristic and the score is not a formal statistical test, it is used as an exploratory ranking measure to prioritize themes for biological interpretation, whereas statistical significance is determined by the adjusted p-values from the underlying ORA and GSEA analyses.

**Supervised tumor subgroup analysis**

Tumor samples annotated as ACC, ACA, or ACX were analyzed using a cell-means limma model (~0 + Subgroup). A moderated F-test (*topTableF*) identified proteins with significant abundance differences across any of the three subgroups (omnibus test). Pairwise follow-up contrasts (ACC vs. ACA, ACC vs. ACX, ACA vs. ACX) were extracted from the same fitted model via *contrasts.fit()*

and *eBayes()*. GSEA on the ACC-versus-ACA contrast specifically used GO:BP (*gseGO*) and Reactome (*gsePathway* via ReactomePA).

**Ordinal trend test.** To detect monotonic abundance trends along a prespecified malignancy axis, subgroup labels were encoded as an ordered numeric covariate (ACA = 1, ACX = 2, ACC = 3) and fitted as a single linear predictor in a limma model (~0 + ordinal). The coefficient for this ordinal term represents the average $\log_2$ abundance change per step along the ACA → ACX → ACC gradient. Proteins with significant positive or negative coefficients (BH-adjusted $P < 0.05$) were reported. As a complementary approach, a pseudo-time projection was derived from PC1 of the tumor-only PCA, and its agreement with the ordinal coding was assessed by Spearman's rank correlation.

**Unsupervised tumor stratification**

Consensus clustering was performed using *ConsensusClusterPlus* on the complete Z-score-normalized protein matrix (all retained proteins, tumor samples only). The full protein feature space was deliberately retained for clustering rather than pre-selecting the most variable features, in order to capture subtle but consistent abundance patterns. Hierarchical clustering was applied using Ward's D2 linkage on Euclidean distances for $k = 2$ through 6, with 1,000 sub-sampling iterations (*pItem* = 0.8, *pFeature* = 1.0), and seed 42. The optimal $k$ was determined by jointly inspecting: (i) the consensus CDF and its relative change in area under the curve (ΔAUC); (ii) the block structure of the consensus matrix at each $k$; and (iii) item-consensus and cluster-consensus statistics computed via *calcICL()*.

As independent validation, a non-negative matrix factorization (NMF) rank survey was performed for ranks 2–6 using the Brunet multiplicative update algorithm (30 runs per rank, seed 42) after shifting the Z-score matrix to non-negative space by subtracting the global minimum value. NMF quality was evaluated using cophenetic correlation, dispersion, and silhouette metrics; a sharp drop in cophenetic correlation was interpreted as evidence of over-splitting beyond the true number of groups. Final cluster assignments were extracted from the consensus matrix at the chosen $k$, and assignment robustness was assessed by silhouette analysis on the consensus distance matrix (1 − consensus).

**Cluster characterization**

**Marker identification.** One-versus-rest limma contrasts were constructed for each cluster: for cluster $C_i$, the contrast was specified as $C_i$ minus the unweighted mean of all remaining clusters. Robust empirical Bayes moderation and BH correction were applied. Proteins with adjusted $P$ <

0.05 and positive $\log_2$ fold change > 0.5 were classified as positive markers for the respective cluster. Only positive markers (elevated abundance in the target cluster) were retained, as these are directly translatable to immunohistochemistry-based detection. The top 20 markers per cluster (ranked by t-statistic) were visualized in annotated heatmaps.

**Clinical associations.** Fisher's exact tests were used for categorical variables (histopathological diagnosis, COG stage, functional activity, endocrine phenotype, sex, vascular invasion, necrosis, DNA methylation group). Kruskal–Wallis tests were used for continuous variables (Ki67 proliferation index, tumor volume, age at diagnosis). Concordance between proteome clusters and the independently determined methylation-based classification was visualized using alluvial diagrams (*ggalluvial*).

**Per-cluster pathway enrichment.** For each cluster, a ranked gene list was constructed from the one-versus-rest limma t-statistics, and GSEA was run against MSigDB Hallmark, C6 oncogenic, and C2:CGP collections. Complementary ORA was performed on cluster-specific increased and decreased proteins against GO:BP, Disease Ontology, and DisGeNET databases. Normalized enrichment scores (NES) were aggregated across all clusters in a summary heatmap to enable systematic cross-cluster comparison of biological programs.

**Overall survival.** OS was measured from diagnosis to death or last follow-up, converted to months (1 month = 30.44 days). Kaplan–Meier curves were constructed for each cluster, and between-group differences were assessed by log-rank tests (*survival* and *survminer* packages). Only tumor samples with non-missing OS time and event indicator were included.

**Marker-based scoring procedure**

Throughout this study, multiple analyses required the computation of per-sample composite scores from curated marker gene panels (adrenal zonation, fetal/adult signatures, immune cell types, IGF2/IGF1R, Wnt/β-catenin, and TP53 pathways). A unified scoring procedure was applied in all cases. For a given marker panel of *n* genes, each gene was first mapped to protein-group identifiers using the annotation table. When multiple protein groups mapped to the same gene symbol, only the one with the highest row-wise variance across all samples was retained, as this maximizes the informative signal-to-noise ratio. The Z-score-normalized abundance value for each detected marker was then extracted. The per-sample composite score was computed as the arithmetic mean of the Z-scores of all detected markers for that panel. A minimum detection threshold was applied (minimum 2 markers required for zonation and immune signatures, minimum 3 for pathway scores); if the number of detected markers fell below this threshold, the score was set to NA. This approach ensures that composite scores are not driven by a single protein measurement and are comparable across panels of different sizes, as Z-score normalization places all proteins on a common scale.

**Adrenal cortical zonation inference**

Curated marker panels for the zona glomerulosa (ZG; 11 markers), zona fasciculata (ZF; 12 markers), zona reticularis (ZR; 9 markers), adrenal medulla (9 markers), and pan-cortical identity (8 markers) were assembled from published zone-specific transcriptomic and immunohistochemical studies (Supplementary Table S1). Using the scoring procedure described above, per-sample zone scores were computed as the mean Z-score of detected markers per zone (minimum 2 detected markers required).

To derive a proportional zone composition suitable for ternary visualization, the raw ZG, ZF, and ZR scores were transformed into proportions via a softmax function. For a given sample with zone scores $s_{ZG}$, $s_{ZF}$, and $s_{ZR}$, the proportion assigned to zone $j$ was computed as:

$$p_j = \exp(s_j) / [\exp(s_{ZG}) + \exp(s_{ZF}) + \exp(s_{ZR})]$$

The softmax transformation converts unbounded Z-score-based scores into non-negative proportions that sum to 1, enabling direct plotting on a ternary composition diagram (*ggtern*). This approach captures the relative zone identity of each sample even when absolute score magnitudes differ. Ternary plots were generated with samples colored by diagnosis (ACA/ACX/ACC) or proteome cluster. Differential zone enrichment across subgroups and clusters was assessed by Kruskal–Wallis rank-sum tests on the raw zone scores, followed by pairwise Wilcoxon rank-sum tests for post-hoc comparisons.

**Fetal versus adult adrenal signature scoring**

To quantify the developmental resemblance of each tumor to fetal versus mature adrenal cortex, curated marker panels were compiled from published studies of human adrenal ontogeny, steroidogenic zonation, and adrenocortical tumorigenesis (Auchus & Rainey, 2004; Drelon et al., 2016; Hui et al., 2009; Lalli & Figueiredo, 2005; Mesiano & Jaffe, 1997; Metherell et al., 2005; Poli et al., 2019, 2024; Rainey & Nakamura, 2008; Rege et al., 2014; Schuetz et al., 1994; Wilkins et al., 2023). Because the present study is based on mass spectrometry-derived protein abundance data, markers were restricted to proteins that are (i) encoded by protein-coding genes and (ii) routinely quantifiable in discovery-grade label-free or DIA proteomics workflows; non-coding RNAs (e.g. H19, MEG3) that are informative in transcriptomic studies were excluded. The fetal adrenal panel comprised 7 proteins enriched in the human fetal adrenal zone: IGF2 (imprinted growth factor, paternal allele), DLK1 (delta-like non-canonical Notch ligand 1), FRZB (secreted frizzled-related protein 3), CYP3A7 (fetal-specific cytochrome P450 3A7), and the DHEA-pathway enzymes CYP17A1 (17alpha-hydroxylase / 17,20-lyase), CYB5A (cytochrome b5, allosteric activator of the CYP17A1 lyase reaction), and SULT2A1 (DHEA sulfotransferase). The inclusion of CYP17A1, CYB5A, and SULT2A1 reflects the zona reticularis-like steroidogenic program of the fetal adrenal, which is dominated by delta-5-steroid production (Rainey et al., 2004). CYP3A7, the predominant CYP3A isoform in fetal liver and adrenal, serves as a fetal-specific marker that is progressively replaced by CYP3A4 and CYP3A5 postnatally.

The adult (mature cortex) panel comprised 8 proteins characteristic of postnatal adrenocortical zonation: STAR (steroidogenic acute regulatory protein; cholesterol import), CYP11A1 (side-chain cleavage; first committed step), HSD3B2 (3beta-hydroxysteroid dehydrogenase type 2; adrenal-specific isoform), CYP21A2 (21-hydroxylase), CYP11B1 (11beta-hydroxylase; zona fasciculata), CYP11B2 (aldosterone synthase; zona glomerulosa), MC2R (ACTH receptor), and MRAP (MC2R accessory protein). These enzymes collectively define the coordinated, ACTH-responsive steroidogenic program of the mature adrenal cortex that is absent or rudimentary in the fetal zone.

Note that three DHEA-pathway genes (CYP17A1, CYB5A, SULT2A1) appear in both the fetal and the zona reticularis signatures. This overlap is biologically expected: the fetal adrenal zone and the postnatal zona reticularis share a common steroidogenic program characterized by low 3-beta-HSD activity and high 17,20-lyase activity, resulting in delta-5-steroid (DHEA/DHEA-S) dominance.

Per-sample fetal and adult scores were computed as the mean z-score of detected panel members (minimum 2 markers required for score calculation). A fetal-to-adult ratio was defined as the arithmetic difference (fetal score minus adult score); positive values indicate a proteome more closely resembling the fetal adrenal, while negative values indicate a mature adrenocortical profile. This ratio was compared between tumor and normal groups by the Wilcoxon rank-sum test and across proteome-derived clusters by the Kruskal-Wallis test.

**Immune microenvironment estimation**

Immune cell infiltration was estimated using curated marker panels for 11 cell types and functional signatures: $CD8^+$ T cells (8 markers), $CD4^+$ T cells (6 markers), regulatory T cells (5 markers), NK cells (6 markers), B cells (6 markers), M1 macrophages (7 markers), M2 macrophages (7 markers), dendritic cells (7 markers), neutrophils (5 markers), mast cells (5 markers), and immune checkpoint molecules (10 markers), compiled from established immune deconvolution signatures (Newman *et al.*, 2015; Bindea *et al.*, 2013; Supplementary Table S6). Per-sample cell-type scores were computed as the mean Z-score of detected markers (minimum 2 required). Scores were compared between tumor and normal tissue and among proteome clusters.

**Kinase activity inference by ssGSEA**

In the absence of phosphoproteomic data, kinase signaling pathway activity was inferred from the abundance levels of pathway members and known substrates using single-sample GSEA (ssGSEA). Kinase-related signaling gene sets were curated from Reactome (CP:REACTOME) and KEGG (CP:KEGG_LEGACY) pathway collections within MSigDB by keyword filtering for terms related to kinase signaling, growth factor receptors, cell cycle, and apoptosis. The ssGSEA score for each gene set in each sample was computed using a rank-based enrichment approach as described by Barbie *et al.* (2009): for each sample, all proteins were ranked by their Z-score, and the enrichment score was calculated as the deviation of the rank-sum of in-set proteins from the expected rank-sum under a uniform distribution, normalized by the product of in-set and out-of-set sizes. A minimum overlap of 5 detected gene-set members was required. Differential kinase pathway activity between tumor and normal tissue and among proteome clusters was assessed by Wilcoxon rank-sum and Kruskal–Wallis tests, respectively.

**Metabolic vulnerability profiling**

Metabolic pathway activity was scored per sample using ssGSEA (as above) on curated gene sets from KEGG metabolic pathways and MSigDB Hallmark metabolic signatures. KEGG metabolic sets were selected by keyword matching for glycolysis, TCA cycle, oxidative phosphorylation, fatty acid and cholesterol metabolism, steroid biosynthesis, amino acid metabolism, nucleotide metabolism, sphingolipid and glycerophospholipid metabolism, xenobiotic metabolism, and peroxisome pathways. Hallmark metabolic sets included glycolysis, oxidative phosphorylation, fatty acid metabolism, cholesterol homeostasis, bile acid metabolism, xenobiotic metabolism, adipogenesis, mTORC1 signaling, reactive oxygen species, peroxisome, and heme metabolism. Differential metabolic pathway activity was assessed between groups and clusters. Tumor-enriched metabolic enzymes were cross-referenced with the druggable-target annotation database to identify exploitable metabolic vulnerabilities.

**Pathway-specific deep-dive analyses**

Three signaling axes of particular relevance to pediatric ACT were profiled in detail using curated gene panels. (i) The IGF2/IGF1R signaling axis (39 genes; Supplementary Table S3) encompassed ligands (IGF1, IGF2, INS), receptors (IGF1R, IGF2R, INSR, INSRR), all seven IGFbinding proteins (IGFBP1–7), and downstream effectors of both the PI3K/AKT/mTOR (14 genes) and RAS/MAPK (11 genes) arms. (ii) The Wnt/β-catenin pathway (41 genes; Supplementary Table S4) covered core signal transduction components, six Wnt ligands, six Frizzled/LRP receptors, TCF/LEF transcription factors, canonical target genes, and a comprehensive set of Wnt modulators and antagonists. (iii) The TP53 pathway (28 genes; Supplementary Table S5) included p53 itself, 17 direct transcriptional targets activated by wild-type p53, 6 negative regulators (MDM2, MDM4, PPM1D, USP7, COP1, PIRH2), and 6 gain-of-function targets associated with mutant p53.

For each pathway, the Z-score abundance of all detected members was extracted and visualized in annotated heatmaps with row-level annotations indicating the functional component (e.g., ligand, receptor, downstream effector). Pathway scores were computed using the marker-based scoring procedure described above. For the TP53 pathway, a composite "p53 functionality score" was additionally derived as the mean Z-score of activated p53 target genes minus the Z-score of TP53 protein itself; high target abundance with low p53 accumulation was interpreted as indicative of wild-type p53, whereas low target abundance with high p53 protein was interpreted as consistent with loss-of-function (mutant) p53.

**Drug repurposing via perturbation-signature matching**

A Connectivity Map (CMap)-equivalent approach was used to identify candidate therapeutic compounds. GSEA was run with the tumor-versus-normal t-statistic-ranked gene list against the full MSigDB C2:CGP collection. Drug-treatment-related gene sets were identified by keyword filtering (e.g., "TREATED", "INHIBIT", compound names). Negatively enriched drug-treatment signatures (NES < 0, BH-adjusted $P < 0.05$) were prioritized as repurposing candidates, as their molecular effects oppose the tumor transcriptional program. The analysis was repeated per proteome cluster using cluster-specific ranked gene lists to identify subtype-selective therapeutic opportunities.

**Druggable proteome annotation**

A curated drug–target annotation database of approximately 360 unique protein targets was compiled from five major drug–target databases: DrugBank, TTD, Open Targets, ChEMBL, and BindingDB (Supplementary Table S2). Each target was annotated with pharmacological family, drug modality (small molecule, monoclonal antibody, ADC, bispecific antibody, CAR-T, PROTAC/molecular glue, peptide, radioligand therapy, trap, or antisense oligonucleotide), cell-surface abundance status, approved drug names, clinical development stage, and source database attribution. Targets were stratified by actionability tier: Tier 1 (FDA-approved), Tier 2 (Phase II/III), Tier 3 (Phase I), Tier 4 (preclinical). The database was intersected with tumor-versus-normal DE results and with cluster-specific marker lists, and summarized in heatmaps of target family and drug modality by cluster.

**Minimal IHC marker panel: leave-one-out cross-validated forward selection**

To derive a compact protein panel for immunohistochemistry (IHC)-based cluster assignment, a greedy forward-selection procedure was implemented as follows.

**Candidate pool.** For each of the *k* proteome clusters, the top 50 significantly increased one-versus-rest markers ($\log_2$ FC > 0.5, BH-adjusted $P < 0.05$) were selected, ranked by the absolute moderated t-statistic. Only positive markers (elevated in the target cluster) were considered, as IHC reliably detects protein overexpression. After de-duplication across clusters (retaining the protein for the cluster in which it showed the strongest effect), the union of all candidates formed the candidate pool.

**Nearest-centroid classifier.** Classification was performed using a Euclidean nearest-centroid algorithm. For a given set of panel proteins, cluster centroids were computed as the mean $\log_2$ abundance profile of all samples assigned to each cluster. A query sample was classified to the cluster whose centroid was nearest in Euclidean distance. This simple parametric classifier was chosen because it is robust with small sample sizes, requires no hyperparameter tuning, and directly mirrors the clinical use case in which a new sample would be compared against cluster-average abundance profiles.

**Leave-one-out cross-validation (LOO-CV).** To obtain an unbiased estimate of classification accuracy, LOO-CV was used. In each of *n* iterations (where *n* is the number of tumor samples), one sample was held out and cluster centroids were recomputed from the remaining $n - 1$ samples. The held-out sample was classified by nearest centroid, and its predicted label was compared to its true consensus-cluster assignment. The LOO-CV accuracy was calculated as the fraction of correctly classified samples across all *n* iterations.

**Forward selection.** Starting from an empty panel, each remaining candidate protein was tentatively added and the LOO-CV accuracy of the resulting panel was computed. The candidate yielding the highest accuracy was permanently added. This greedy process was repeated iteratively up to a maximum panel size of 20 proteins or until 100% LOO-CV accuracy was achieved. The accuracy was recorded at each step to produce an accuracy-versus-panel-size trace.

**Panel size selection.** The optimal panel size was determined as the smallest number of proteins at which the LOO-CV accuracy reached at least 95% of the maximum observed accuracy across all panel sizes, subject to a minimum panel size of 5 proteins. This parsimony criterion avoids overfitting to a particular panel size and favors a smaller number of IHC stains for practical diagnostic applicability.

**Permutation test.** Statistical significance of the observed LOO-CV accuracy was assessed by a label-permutation test. Cluster labels were randomly shuffled 1,000 times, and for each permutation the LOO-CV accuracy of the selected panel was recomputed. The empirical *P*-value was calculated as $(m + 1) / (N + 1)$, where $m$ is the number of permutations achieving accuracy greater than or equal to the observed accuracy and $N = 1{,}000$ is the total number of permutations. The addition of 1 to both numerator and denominator provides a conservative correction.

**Supplemental Tables**

**Supplementary Table S1. Demographic and clinical characteristics of 72 children and adolescents with adrenocortical tumors (pACT).**

| Characteristic | Subcategory | Patients (n=72) | % |
|---|---|---|---|
| **Diagnosis** | ACC | 36 | 50.0 |
| | ACA | 24 | 33.3 |
| | ACx | 12 | 16.7 |
| **Age at diagnosis (years)** | 0–4 | 39 | 54.2 |
| | >4–10 | 14 | 19.4 |
| | >10–18 | 19 | 26.4 |
| **Gender** | Male | 22 | 30.6 |
| | Female | 50 | 69.4 |
| **Endocrine phenotype** | Virilization/Precocious Puberty | 47 | 65.3 |
| | Cushing's syndrome | 19 | 26.4 |
| | Mixed | 12 | 16.7 |
| | Silent | 17 | 23.6 |
| | Unknown | 1 | 1.4 |
| **Ki67 index** | 0–10% | 34 | 47.2 |
| | >10–20% | 15 | 20.8 |
| | >20% | 21 | 29.2 |
| | Unknown | 2 | 2.8 |
| **COG Stage** | 1 | 10 | 13.9 |
| | 2 | 9 | 12.5 |
| | 3 | 20 | 27.8 |
| | 4 | 6 | 8.3 |
| | Unknown | 3 | 4.2 |

| | | | |
|---|---|---|---|
| **pT Stage** | T1 | 19 | 26.4 |
| | T2 | 34 | 47.2 |
| | T3 | 1 | 1.4 |
| | T4 | 14 | 19.4 |
| | Unknown | 4 | 5.6 |
| **Status at last follow-up** | Dead of disease | 11 | 15.3 |
| | Alive in complete remission | 54 | 75.0 |
| | Alive with disease | 2 | 2.8 |
| | Unknown | 5 | 6.9 |

**Supplementary Table S2. Curated adrenal cortical zone marker panels**

References: Vidal *et al.* 2016; Drelon *et al.* 2016; Nishimoto *et al.* 2015; Rege *et al.* 2014, 2022; Altieri *et al.* 2024; Auchus *et al.* 1998; Pihlajoki *et al.* 2015; Gorrigan *et al.* 2011; Bassett *et al.* 2004; Zanaria *et al.* 1994.

| Zone | Protein | Description / rationale |
|---|---|---|
| ZG | CYP11B2 | Aldosterone synthase; definitive ZG marker |
| ZG | DAB2 | Disabled-2; Wnt target, ZG-enriched |
| ZG | SHH | Sonic Hedgehog; ZG progenitor niche |
| ZG | NRCAM | Neuronal cell adhesion molecule; ZG-specific |
| ZG | NRP1 | Neuropilin-1; ZG-enriched |
| ZG | VSNL1 | Visinin-like 1; IHC-validated ZG (Nishimoto 2015) |
| ZG | WNT4 | Wnt ligand; ZG-enriched (Vidal 2016) |
| ZG | DACH1 | Dachshund 1; subcapsular/ZG (Altieri et al. 2024) |
| ZG | AGTR1 | Angiotensin II type 1 receptor |
| ZG | NPNT | Nephronectin; ZG-enriched (spatial transcriptomics) |
| ZG | NCAM1 | Neural cell adhesion molecule; ZG regenerative cells |
| ZF | CYP11B1 | 11β-hydroxylase; definitive ZF marker |
| ZF | CYP21A2 | 21-hydroxylase; cortisol pathway |
| ZF | CYP17A1 | 17α-hydroxylase; shared ZF/ZR, high in ZF |
| ZF | MC2R | ACTH receptor (Pihlajoki et al. 2015) |
| ZF | MRAP | MC2R accessory protein (Gorrigan et al. 2011) |
| ZF | LEF1 | Wnt TF; most upregulated ZF vs. ZR (Rege et al. 2014) |
| ZF | NOV | CCN3; ZF-enriched (Rege et al. 2014) |
| ZF | PCP4 | Purkinje cell protein 4; ZF-enriched (Rege et al. 2014) |
| ZF | HSD3B2 | 3β-HSD type 2; ZG + ZF, absent ZR |
| ZF | SCARB1 | SR-B1; HDL cholesterol receptor |
| ZF | FDX1 | Ferredoxin 1; mitochondrial electron transfer |

| | | |
|---|---|---|
| ZF | LDLR | LDL receptor; cholesterol uptake |
| ZR | CYB5A | Cytochrome b5; definitive ZR marker (Auchus 1998) |
| ZR | SULT2A1 | DHEA sulfotransferase |
| ZR | AKR1C3 | Type 5 17β-HSD; androgen metabolism |
| ZR | PAPSS2 | PAPS synthase 2; required for SULT2A1 (Noordam et al. 2009) |
| ZR | SLC27A2 | FATP2; ZR top transcript (Rege et al. 2014) |
| ZR | TSPAN12 | Tetraspanin 12; IHC-validated ZR (Rege et al. 2014) |
| ZR | FGG | Fibrinogen-γ; most upregulated ZR transcript (Rege et al. 2014) |
| ZR | GSTA1 | GST-A1; ZR pigmentation/metabolism |
| ZR | SRD5A1 | 5α-reductase type 1; androgen metabolism |
| Medulla | TH | Tyrosine hydroxylase; rate-limiting catecholamine synthesis |
| Medulla | DBH | Dopamine β-hydroxylase |
| Medulla | PNMT | Phenylethanolamine N-methyltransferase |
| Medulla | CHGA | Chromogranin A |
| Medulla | CHGB | Chromogranin B |
| Medulla | DDC | DOPA decarboxylase |
| Medulla | SLC18A1 | VMAT1; vesicular monoamine transporter |
| Medulla | SYP | Synaptophysin |
| Medulla | SCG2 | Secretogranin II |
| Pan-cortical | STAR | StAR; cholesterol import to mitochondria |
| Pan-cortical | CYP11A1 | Cholesterol side-chain cleavage (P450scc) |
| Pan-cortical | HSD3B2 | 3β-HSD type 2 (ZG + ZF) |
| Pan-cortical | NR5A1 | SF-1; master steroidogenic TF |
| Pan-cortical | NR0B1 | DAX1; adrenal development (Zanaria et al. 1994) |
| Pan-cortical | POR | P450 oxidoreductase |
| Pan-cortical | FDXR | Ferredoxin reductase |

| | | |
|---|---|---|
| Pan-cortical | SCARB1 | SR-B1; HDL cholesterol uptake |

**Supplementary Table S3. Druggable proteome: pharmacological target families**

Sources: DrugBank, TTD, Open Targets, ChEMBL, BindingDB. ~360 unique targets.

| Family | n | Key members |
|---|---|---|
| Receptor tyrosine kinases | 33 | EGFR, ERBB2/3/4, FGFR1–4, PDGFRA/B, KIT, VEGFR1–3, FLT3, MET, AXL, RET, ALK, ROS1, IGF1R, INSR, NTRK1–3, EPH receptors, DDR1/2, TEK, CSF1R |
| Non-receptor kinases | 57 | ABL1/2, SRC, JAK1/2/3/TYK2, RAF/MEK/ERK, CDK4/6/1/2/7/9, Aurora A/B, PLK1/4, CHK1/2, WEE1, ATR/ATM, mTOR, PI3Kα/β/δ/γ, AKT1–3, GSK3B, ROCK1/2 |
| GPCRs | 35 | Adenosine, adrenergic, muscarinic, dopamine, serotonin, S1P, chemokine, SMO, FZD7, GLP1R, SSTR2/5 |
| Ion channels | 22 | $Na^+$/$K^+$/$Ca^{2+}$ voltage-gated, AMPA/NMDA/GABA ligand-gated, TRP (V1/4, M8), CHRNA7, P2X7, CFTR |
| Nuclear receptors | 22 | ESR1/2, AR, GR, MR, PPARα/γ/δ, VDR, RAR, RXR, FXR, LXR, THRα/β, PGR |
| Proteases | 23 | MMPs, ADAMs, cathepsins, caspases, DUBs, thrombin, FXa, DPP4, ACE/ACE2, TMPRSS2 |
| Phosphatases | 8 | PTPN11/1/2, PP2A, DUSP1/6, CDC25A/C |
| Phosphodiesterases | 6 | PDE4A/B/D, PDE5A, PDE3A, PDE10A |
| Metabolic enzymes | 32 | COX-1/2, HMGCR, FASN, ACLY, DHFR, TYMS, DHODH, PKM, LDHA/B, HK2, GLS, NAMPT, IDO1 |
| Epigenetic regulators | 32 | HDAC1–11, SIRT1/2, EZH2, DNMT1/3A/3B, BRD2/3/4, KDM1A/5A/B, PRMT1/5, IDH1/2 |
| Chaperones | 6 | HSP90AA1/AB1/B1, HSPA5, HSPA4, TRAP1 |
| Cytoskeletal | 6 | TUBB/TUBB3, TUBA1A/B, KIF11, DYNC1H1 |
| DNA/RNA processing | 12 | TOP1, TOP2A/B, PARP1/2/3, POLA1, WRN, SF3B1, DDX3X |
| Immune checkpoints | 12 | CD274, PDCD1, CTLA4, LAG3, HAVCR2, TIGIT, CD47, SIRPA, TNFRSF9, ICOS |

| | | |
|---|---|---|
| Surface targets | 23 | NECTIN4, TROP2, CEACAM5, FOLR1, MSLN, DLL3, CD276, EPCAM, GPC3, CLDN18/6, MUC1/16 |
| Growth factors/ligands | 22 | VEGFA/B/C, TNF, IL-6, IL-1, TGFβ1/2, EGF, HGF, IGF1/2, ANGPT2, RANKL |
| Proteasome/apoptosis | 15 | PSMB5/8/9, BCL2, BCL-XL, MCL1, XIAP, survivin, MDM2/4, CRBN |
| Transporters | 13 | ABCB1 (P-gp), ABCG2, ABCC1, SLC7A11, SLC7A5, SLC2A1, SLC6A4, ATP1A1 |
| Signalling hubs | 23 | TP53, CTNNB1, NOTCH1/2, KRAS/NRAS, MYC/MYCN, HIF1α/2α, NRF2, STAT3, TERT, YAP1 |
| ACT-relevant | 12 | CYP11A1, CYP11B1/B2, CYP17A1, CYP19A1, CYP21A2, HSD3B2, STAR, NR5A1, NR0B1, MC2R, SULT2A1 |

**Supplementary Table S4. IGF2/IGF1R signaling pathway panel (39 proteins)**

| Component | Proteins |
|---|---|
| Ligands (3) | IGF2, IGF1, INS |
| Receptors (4) | IGF1R, IGF2R, INSR, INSRR |
| Binding proteins (7) | IGFBP1, IGFBP2, IGFBP3, IGFBP4, IGFBP5, IGFBP6, IGFBP7 |
| PI3K/AKT/mTOR arm (14) | IRS1, IRS2, PIK3CA, PIK3CB, PIK3R1, AKT1, AKT2, AKT3, MTOR, RPS6KB1, EIF4EBP1, PTEN, TSC1, TSC2 |
| RAS/MAPK arm (11) | GRB2, SOS1, HRAS, KRAS, NRAS, RAF1, BRAF, MAP2K1, MAP2K2, MAPK1, MAPK3 |

**Supplementary Table S5. Wnt/β-catenin pathway panel (41 proteins)**

| Component | Proteins |
|---|---|
| Core pathway (8) | CTNNB1, APC, AXIN1, AXIN2, GSK3B, DVL1, DVL2, DVL3 |
| Ligands (6) | WNT4, WNT2B, WNT3, WNT5A, WNT7A, WNT11 |
| Receptors (6) | FZD1, FZD2, FZD5, FZD7, LRP5, LRP6 |
| Transcription factors (4) | LEF1, TCF7, TCF7L1, TCF7L2 |
| Canonical targets (3) | MYC, CCND1, AXIN2 |
| Modulators/antagonists (10) | RSPO3, RNF43, ZNRF3, DKK1, DKK3, SFRP1, SFRP2, NKD1, NKD2, NOTUM |

**Supplementary Table S6. TP53 pathway panel (28 proteins)**

| Component | Proteins |
|---|---|
| Core (1) | TP53 |
| Activated targets (17) | CDKN1A, MDM2, BAX, BBC3, PMAIP1, GADD45A, GADD45B, DDB2, XPC, SESN1, SESN2, TIGAR, FAS, PERP, TP53I3, RRM2B, SFN |
| Negative regulators (6) | MDM2, MDM4, PPM1D, USP7, COP1 (RFWD2), PIRH2 (RCHY1) |
| Gain-of-function targets (6) | MYC, VEGFA, PCNA, EGR1, STMN1, PIN1 |

**Supplementary Table S7. Immune cell-type marker panels**

Based on Newman *et al.* 2015 (CIBERSORT) and Bindea *et al.* 2013.

| Cell type (n markers) | Marker proteins |
|---|---|
| CD8+ T cells (8) | CD8A, CD8B, GZMA, GZMB, GZMK, PRF1, IFNG, NKG7 |
| CD4+ T cells (6) | CD4, IL7R, CCR7, SELL, LEF1, TCF7 |
| Regulatory T cells (5) | FOXP3, IL2RA, CTLA4, IKZF2, TNFRSF18 |
| NK cells (6) | NCAM1, KLRB1, KLRD1, KLRK1, NKG7, GNLY |
| B cells (6) | CD19, MS4A1, CD79A, CD79B, BANK1, PAX5 |
| Macrophages M1 (7) | CD68, CD80, CD86, NOS2, IL1B, TNF, CXCL10 |
| Macrophages M2 (7) | CD68, CD163, MRC1, MSR1, CD209, IL10, TGFB1 |
| Dendritic cells (7) | ITGAX, CD1C, CLEC10A, FCER1A, HLA-DRA, HLA-DRB1, HLA-DPA1 |
| Neutrophils (5) | FCGR3B, CEACAM8, CSF3R, CXCR1, CXCR2 |
| Mast cells (5) | TPSAB1, TPSB2, CPA3, KIT, MS4A2 |
| Immune checkpoints (10) | CD274, PDCD1LG2, PDCD1, CTLA4, LAG3, HAVCR2, TIGIT, IDO1, CD47, SIGLEC15 |

**Supplementary Table S8. Fetal and adult adrenal cortex marker panels**

Based on Auchus & Rainey 2004; Drelon *et al*. 2016; Hui *et al*. 2009; Lalli & Figueiredo 2005; Mesiano & Jaffe 1997; Metherell *et al*. 2005; Poli *et al*. 2019, 2024; Rainey & Nakamura 2008; Rege *et al*. 2014; Schuetz *et al*. 1994; Wilkins *et al*. 2023.

| Signature | Protein | Description |
|---|---|---|
| Fetal | IGF2 | Imprinted growth factor; overexpressed in fetal zone and pediatric ACT |
| Fetal | DLK1 | Delta-like 1; fetal zone marker (Poli et al.) |
| Fetal | FRZB | SFRP3; Wnt modulator, fetal zone |
| Fetal | SFRP1 | Secreted frizzled-related protein 1 |
| Fetal | CYP3A7 | fetal-specific cytochrome P450 3A7 |
| Fetal | CYP17A1 | 17α-hydroxylase; high in fetal zone (DHEA pathway) |
| Fetal | CYB5A | Cytochrome b5a; CYP17A1 lyase activator |
| Fetal | SULT2A1 | DHEA sulfotransferase |
| Adult | HSD3B2 | 3β-HSD2; definitive cortex (ZG + ZF) |
| Adult | CYP11B1 | 11β-hydroxylase; ZF cortisol synthesis |
| Adult | CYP11B2 | Aldosterone synthase; ZG-specific |
| Adult | CYP21A2 | 21-hydroxylase; all cortical zones |
| Adult | MC2R | ACTH receptor |
| Adult | MRAP | MC2R accessory protein |
| Adult | STAR | StAR; mature steroidogenic cells |
| Adult | CYP11A1 | Cholesterol side-chain cleavage; pan-cortical |

**Supplementary Table S9. Software, R and Bioconductor packages**

All analyses were performed in R v4.5.2 (R Core Team, 2025) with Bioconductor v3.22 (Huber *et al.*, *Nature Methods*, 2015).

| Package | Version | Citation |
|---|---|---|
| *dplyr* | 1.2.0 | Wickham H et al. (2023). dplyr: A Grammar of Data Manipulation. R package. |
| *tidyr* | 1.3.2 | Wickham H et al. (2024). tidyr: Tidy Messy Data. R package. |
| *ggplot2* | 4.0.2 | Wickham H (2016). ggplot2: Elegant Graphics for Data Analysis. Springer. |
| *ggrepel* | 0.9.6 | Slowikowski K (2024). ggrepel: Auto-Positioning Non-Overlapping Text Labels. R package. |
| *limma* | 3.66.0 | Ritchie ME et al. (2015). Nucleic Acids Res 43(7):e47. |
| *pheatmap* | 1.0.13 | Kolde R (2019). pheatmap: Pretty Heatmaps. R package. |
| *umap* | 0.2.10.0 | Konopka T (2023). umap: Uniform Manifold Approximation and Projection. R package. |
| *clusterProfiler* | 4.18.4 | Wu T et al. (2021). Innovation 2(3):100141. |
| *org.Hs.eg.db* | 3.22.0 | Carlson M (2024). org.Hs.eg.db: Genome wide annotation for Human. Bioconductor. |
| *RColorBrewer* | 1.1-3 | Neuwirth E (2022). RColorBrewer: ColorBrewer Palettes. R package. |
| *ReactomePA* | 1.54.0 | Yu G & He QY (2016). Mol BioSyst 12(2):477–479. |
| *enrichplot* | 1.30.4 | Yu G (2024). enrichplot: Visualization of Functional Enrichment Result. Bioconductor. |
| *ggtext* | 0.1.2 | Wilke CO (2022). ggtext: Improved Text Rendering for ggplot2. R package. |
| *ggnewscale* | 0.5.2 | Campitelli E (2024). ggnewscale: Multiple Fill and Colour Scales in ggplot2. R package. |
| *vegan* | 2.7-2 | Oksanen J et al. (2024). vegan: Community Ecology Package. R package. |

| | | |
|---|---|---|
| *ggtern* | 4.0.0 | Hamilton NE & Ferry M (2018). J Stat Softw 87(3):1–17. |
| *msigdbr* | 25.1.1 | Dolgalev I (2024). msigdbr: MSigDB Gene Sets for Multiple Organisms. R package. |
| *DOSE* | 4.4.0 | Yu G et al. (2015). Bioinformatics 31(4):608–609. |
| *ConsensusClusterPlus* | 1.74.0 | Wilkerson MD & Hayes DN (2010). Bioinformatics 26(12):1572–1573. |
| *NMF* | 0.28 | Gaujoux R & Seoighe C (2010). BMC Bioinformatics 11:367. |
| *cluster* | 2.1.8.2 | Maechler M et al. (2024). cluster: Finding Groups in Data. R package. |
| *survival* | 3.8-6 | Therneau TM (2024). A Package for Survival Analysis in R. R package. |
| *survminer* | 0.5.1 | Kassambara A et al. (2021). survminer: Survival Curves. R package. |
| *ggalluvial* | 0.12.5 | Brunson JC (2020). R J 12(1):7–28. |
| *glmnet* | 4.1-10 | Friedman JH et al. (2010). J Stat Softw 33(1):1–22. |
